# Factors Affecting the Implementation of Continuous Positive Airway Pressure (CPAP) in Low- and Middle-Income Countries: A Qualitative Evidence Synthesis

**DOI:** 10.1101/2025.04.13.25325736

**Authors:** Sindhu Sivanandan, Ambalakkuthan Murugesan, Monisha Rameshbabu, M Jeeva Sankar

## Abstract

**Objective:** To identify the facilitators and barriers affecting the implementation of CPAP in low- and middle-income countries (LMICs).

**Methods:** Electronic databases—PubMed, Embase, Cochrane CENTRAL, EMBASE, and EBSCO—were searched from their inception until July 2, 2023. All types of studies reporting factors that influence CPAP implementation in neonates with respiratory distress were included. The review adhered to the Preferred Reporting Items for Systematic Reviews and Meta-Analyses (PRISMA) guidelines and was registered with PROSPERO (CRD42024497038). The primary outcomes were the facilitators and barriers to CPAP implementation. The identified facilitators and barriers were categorized into five key themes, along with various subthemes under each: medical devices (intrinsic characteristics of CPAP and cost), service delivery (availability of equipment, infrastructure, supplies, and accessories, and monitoring devices), workforce (availability of staff and training), information (perceptions of parents, healthcare workers, and communication), and governance and leadership.

**Findings:** Of the 42 studies included in the review, 30 were conducted in Africa, 9 in Asia, 1 each from Oceania and Central America, and 1 across multiple LMICs; 15 were multi-center studies. Among the facilitators, the most frequently identified subthemes, listed in decreasing order of frequency, were the training of health personnel (n=23), the availability of structured unit protocols for CPAP administration (n=12), and the ease of use of the CPAP device (n=10). The main barriers identified included staff shortages (n=15), a lack of CPAP devices (n=11), and insufficient supplies and accessories for CPAP administration (n=10).

**Conclusions:** Sufficient staffing, ongoing training, and appropriate infrastructure are essential for the effective implementation of CPAP in LMIC settings.

## Introduction

Continuous positive airway pressure (CPAP) is used to prevent and treat respiratory distress in neonates. It reduces the need for invasive ventilation, surfactant therapy, bronchopulmonary dysplasia, and mortality (1,2). Despite its proven benefits, the implementation of CPAP in low- and middle-income countries (LMICs) remains suboptimal (3). Kinsella et al. observed that the ease of use, affordability, and simple maintenance of bubble CPAPs, along with staff training, facilitated its adoption in Sub-Saharan Africa (4). They identified significant barriers, including staff shortages, a lack of CPAP devices, inadequate infrastructure, and caregiver apprehension. Dada et al. (5) reported similar findings regarding the facilitators and obstacles to CPAP implementation in 19 countries. There is limited evidence from LMICs outside of Africa on factors affecting CPAP implementation; therefore, this systematic review was designed to provide updated evidence on the facilitators and barriers to CPAP use in neonates across all LMICs.

## Methods

### Inclusion and exclusion criteria

This review included randomized controlled trials, quasi-experimental studies, observational and exploratory studies, qualitative studies, economic evaluations, program reports, and quality improvement studies that addressed the implementation and utilization of CPAP systems among neonates (≤[28[days old). We included studies conducted at health facilities in LMICs as classified by the World Bank (6). We excluded studies that enrolled children where the neonatal representation was less than one-third, studies conducted in delivery rooms or emergency departments, studies reporting other forms of non-invasive support such as non-invasive positive pressure ventilation or heated humidified nasal cannula therapy, studies presented as abstracts with incomplete data, bench-top studies, literature reviews, letters to the editor, opinion pieces, editorials, and studies published in languages other than English.

### Search strategy

We systematically reviewed relevant publications by searching electronic databases from their inception until July 2, 2023: MEDLINE (1966-July 2023) via PubMed, Cochrane Central Register of Controlled Trials (CENTRAL, The Cochrane Library, Issue 6, June 2023), EMBASE (1988-July 2, 2023), CINAHL (1981-July 2, 2023), and EBSCO databases. We employed the search terms “continuous positive airway pressure,” “CPAP,” “positive distending pressure,” and “neonates OR infants” in our search strategy (see **Appendix)**. Additionally, we examined the clinical trial databases and the reference lists of retrieved articles for eligible studies.

### Outcomes

The critical outcomes were the facilitators and barriers to implementing or scaling up CPAP in newborn units in LMIC settings.

### Selection of studies

We compiled records from various sources into a single database (Covidence) and eliminated duplicates. Three review authors (SS, MA, MS) independently evaluated the titles and abstracts of the identified records to determine their potential eligibility. We retrieved the full text of all articles deemed likely to be relevant. Two review authors assessed these articles independently based on the inclusion and exclusion criteria. We resolved any disagreements through discussion or, if needed, by consulting the fourth author’s (MJS) opinion. Each study was independently evaluated by two review authors who extracted data using an e-form piloted in three studies. The third author verified the data extraction for accuracy and completeness. We extracted the following information from each study: study method, sample size, country, facility type, CPAP type, outcomes, complications, barriers, and facilitators.

### Strategy for data synthesis

Since the review focused on the qualitative synthesis of facilitators and barriers to CPAP implementation, we did not conduct a quality assessment of the included studies. The individual facilitators and barriers were ranked in order of importance based on the number of studies that reported each.

## Results

The number of articles identified from the different databases and the PRISMA flow chart is provided in Figure 1. A total of 42 studies were identified (7–48). Table 1 provides the study characteristics, including the country of origin and the study design. Thirty were from the African continent (7–10, 12–16, 20, 21, 23, 24, 26, 27, 32–40, 42–46), nine were from Asia (11, 17–19, 22, 25, 29–31), one each from Fiji (28) and Central America (41), while one study involved countries from different continents (48). While most were single-center studies, 13 and 2 studies from Africa and Asia, respectively, included multiple centers. All the studies were conducted between 2003 and 2022, with most (n=33) performed in the last decade. Twenty-eight studies reported the clinical application of CPAP in neonates, 12 reported healthcare workers’ perceptions, and 2 reported parental perceptions. The study designs included prospective observational, pre-post interventional studies, qualitative studies, quality improvement, cross-sectional descriptive, and randomized controlled studies (Table 1).

**Figure 1:**
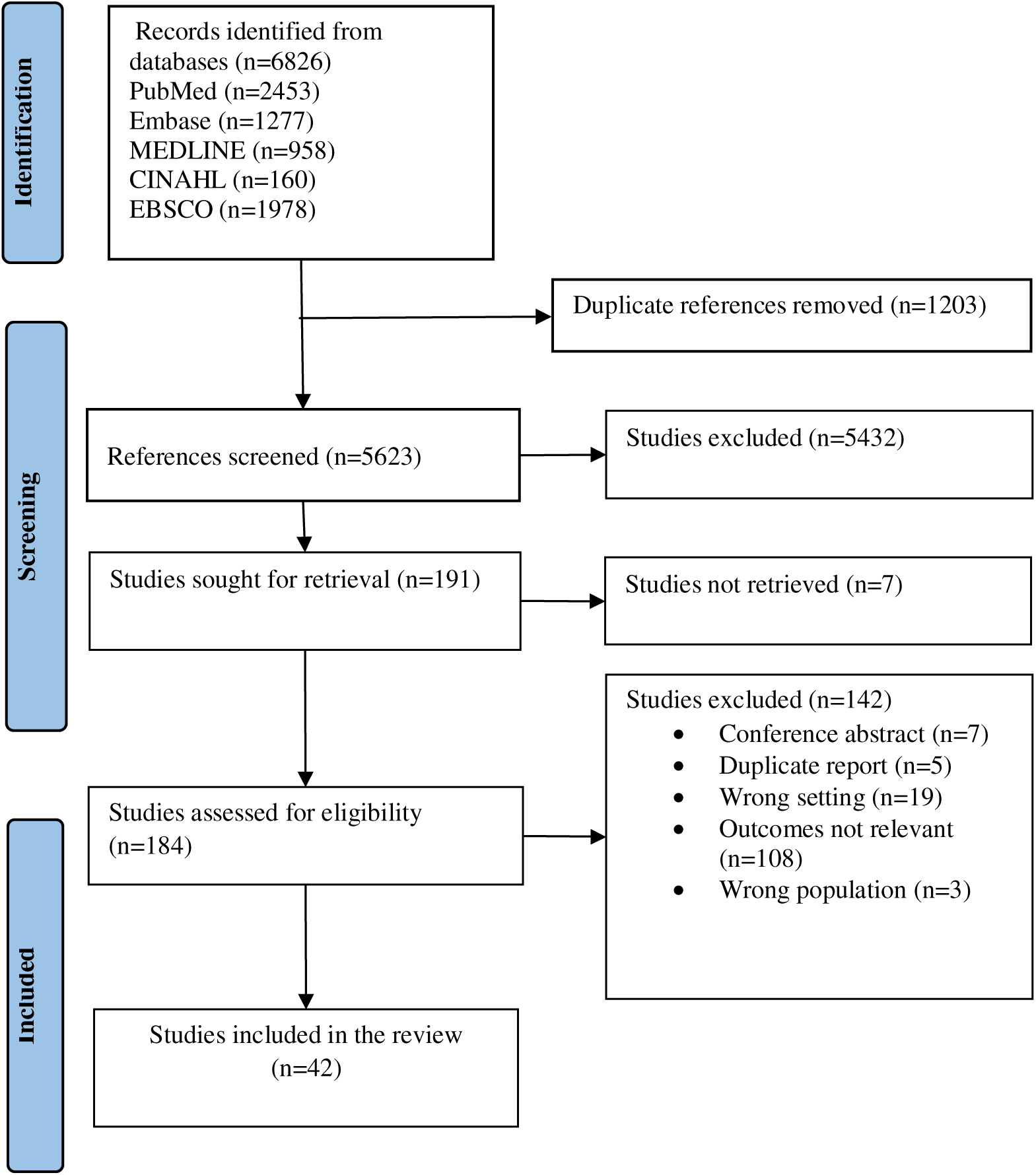
PRISMA flow diagram

**Table 1:** Characteristics of included studies.

| **Study characteristics** | **No. of studies** |
| --- | --- |
| **Countries** |  |
| **Africa** | **30** |
| Malawi | 14 |
| Kenya | 5 |
| Nigeria | 2 |
| Uganda | 3 |
| Ghana | 1 |
| Tanzania | 2 |
| Burkina Faso | 1 |
| Ethiopia | 1 |
| Multiple countries in Africa | 1 |
| **Asia** | **9** |
| India | 5 |
| Pakistan | 2 |
| Iraq | 1 |
| Armenia | 1 |
| **Oceania** | **1** |
| Fiji |  |
| **Central America** | **1** |
| Nicaragua |  |
| Multiple continents (5 LMIC – Ghana, Cambodia, Honduras, Kenya, Rwanda) | **1** |
| **Study design** |  |
| Prospective observational study | 7 |
| Pre-post intervention study | 8 |
| Qualitative study | 7 |
| Quality improvement study | 5 |
| Cross-sectional study | 5 |
| Descriptive study (questionnaire – 2, retrospective chart review – 1, description of CPAP training programme – 1) | 4 |
| Randomized controlled trial | 3 |
| Cost-effectiveness study | 2 |
| Retrospective cohort | 1 |

The type of CPAP device used was mentioned in 33 studies. The Pumani CPAP device was used in 13 studies (8, 10, 13–16, 26, 27, 33, 36), followed by the Diamedica device in three studies (9, 37, 38), and the PATH CPAP (23), DeVilbiss IntelliPAP (39), Vayu CPAP (45), and MTTS CPAP (40) in one study each. A bubble CPAP device with a simple oxygen flow meter, nasal prongs, and a bubble chamber made from locally available material was used in six studies (12, 19, 20, 25, 32, 34, 47). Two studies (21, 24) did not mention the CPAP type; another six (17, 18, 35, 43, 46, 48) focussed on caregiver or healthcare worker perceptions and CPAP availability and did not mention the CPAP device characteristics.

### Facilitators and barriers to CPAP implementation

The facilitators and barriers identified from the studies were categorized into the following themes: those related to medical devices (intrinsic characteristics of CPAP), service delivery (availability of equipment, infrastructure, supplies, and accessories, monitoring devices), workforce (availability of staff and training), information (perceptions of caregivers and healthcare workers, as well as communication), and governance and leadership (as shown in Figure 2 and Table 6 in the Appendix). Table 2 summarizes the facilitators and barriers reported in the included studies.

**Figure 2:**
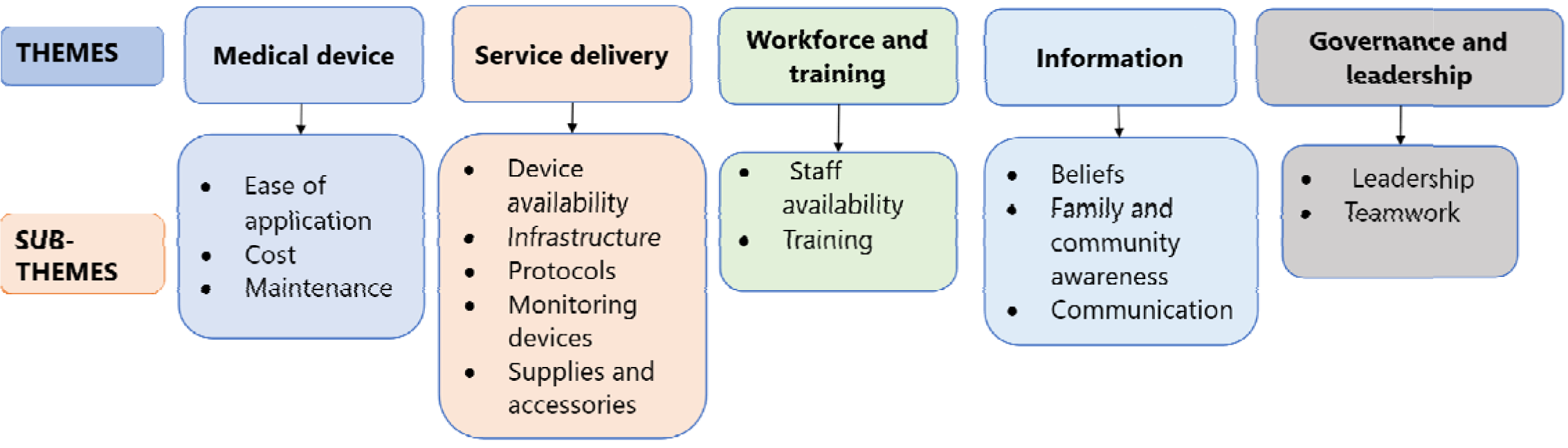
Facilitators and barriers under various themes and subthemes

**Table 2:**
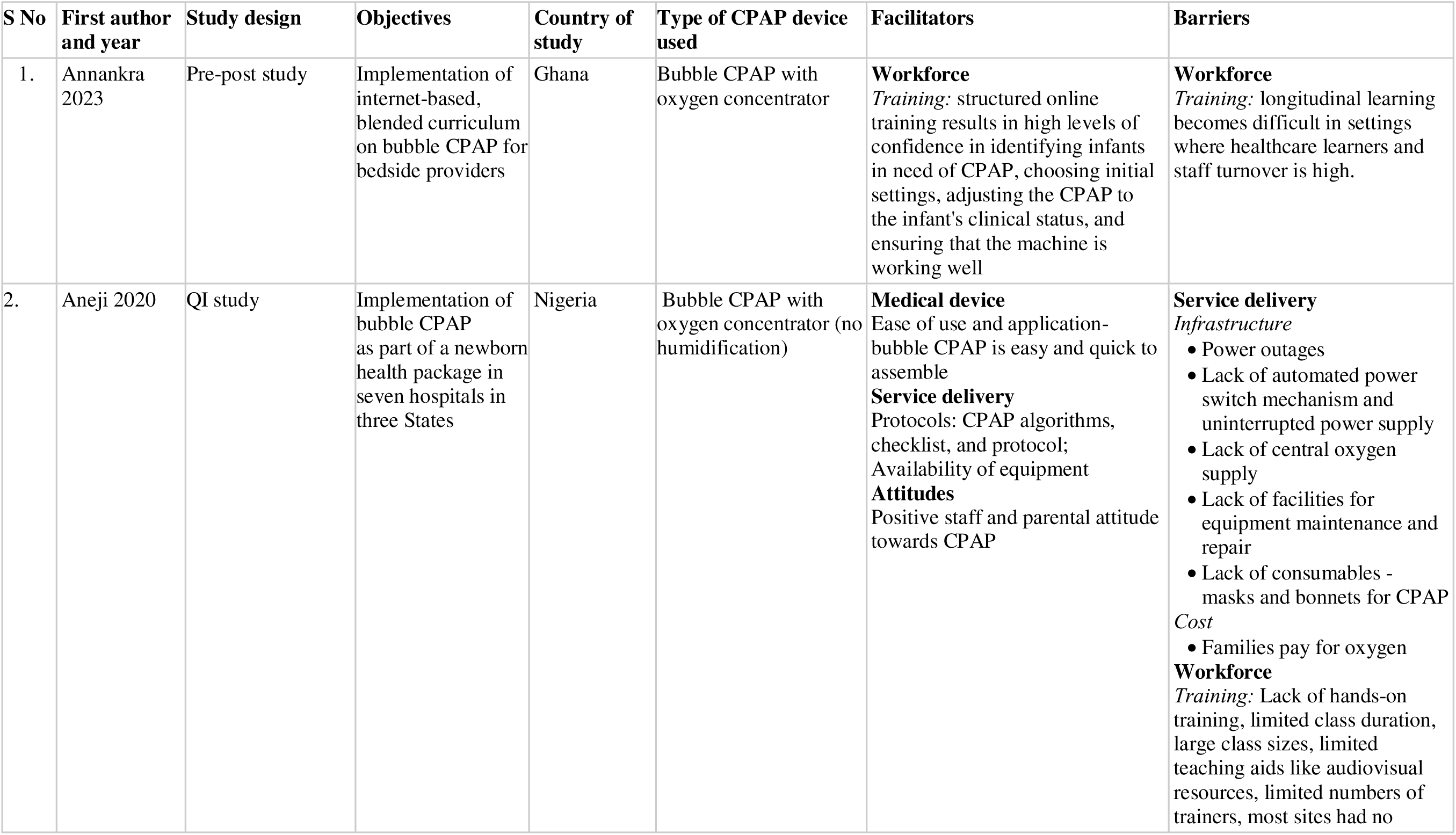
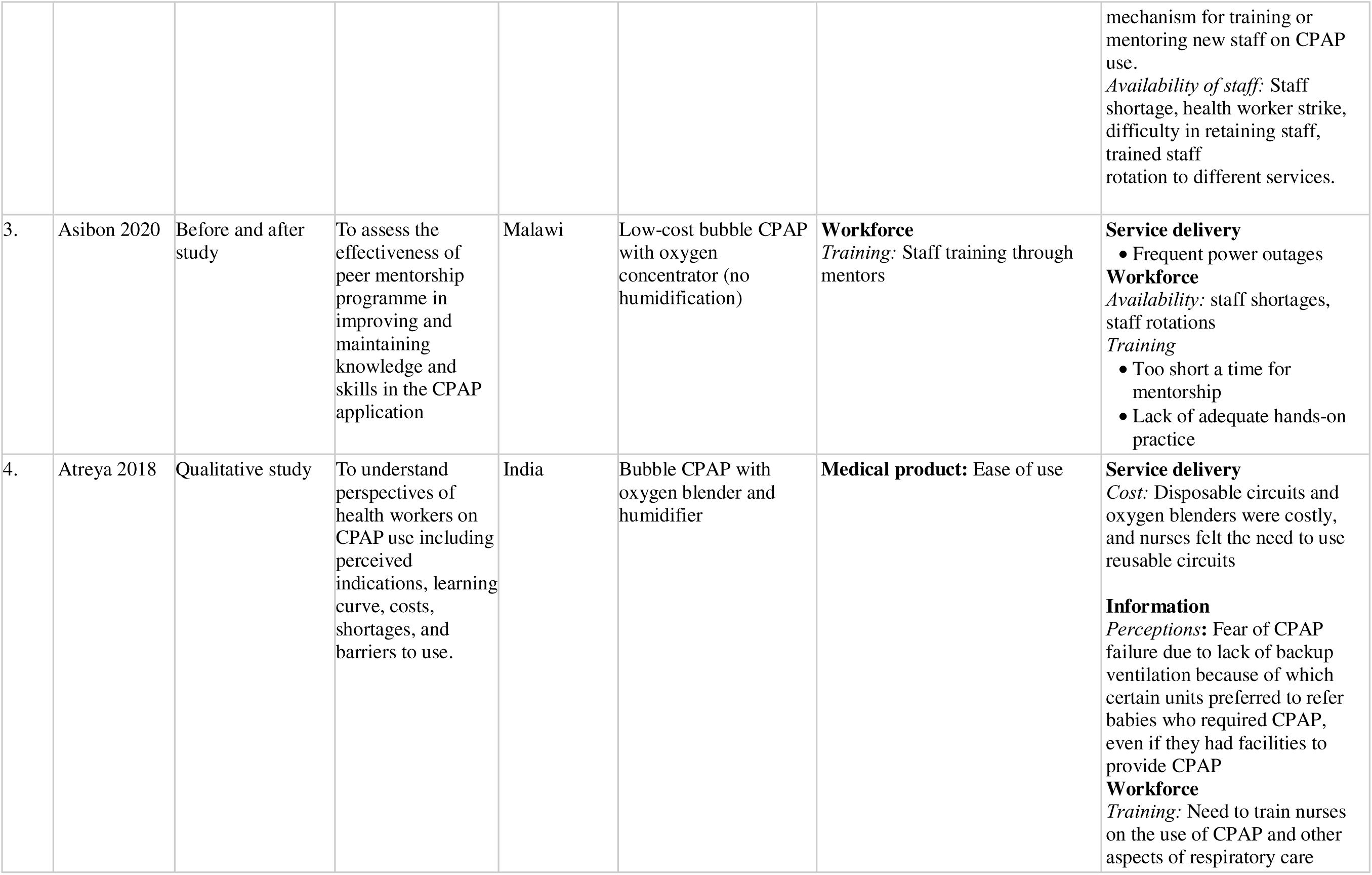
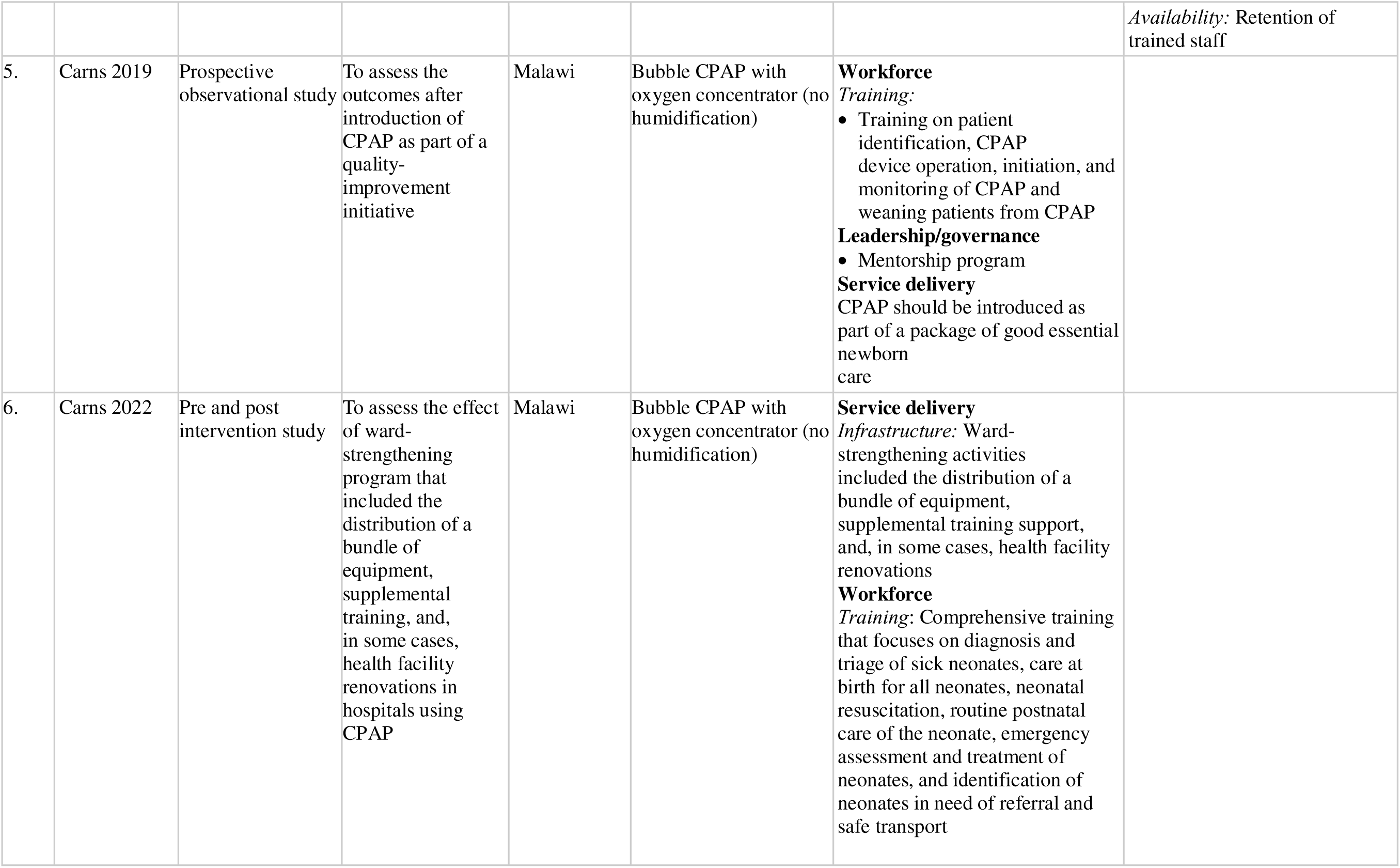
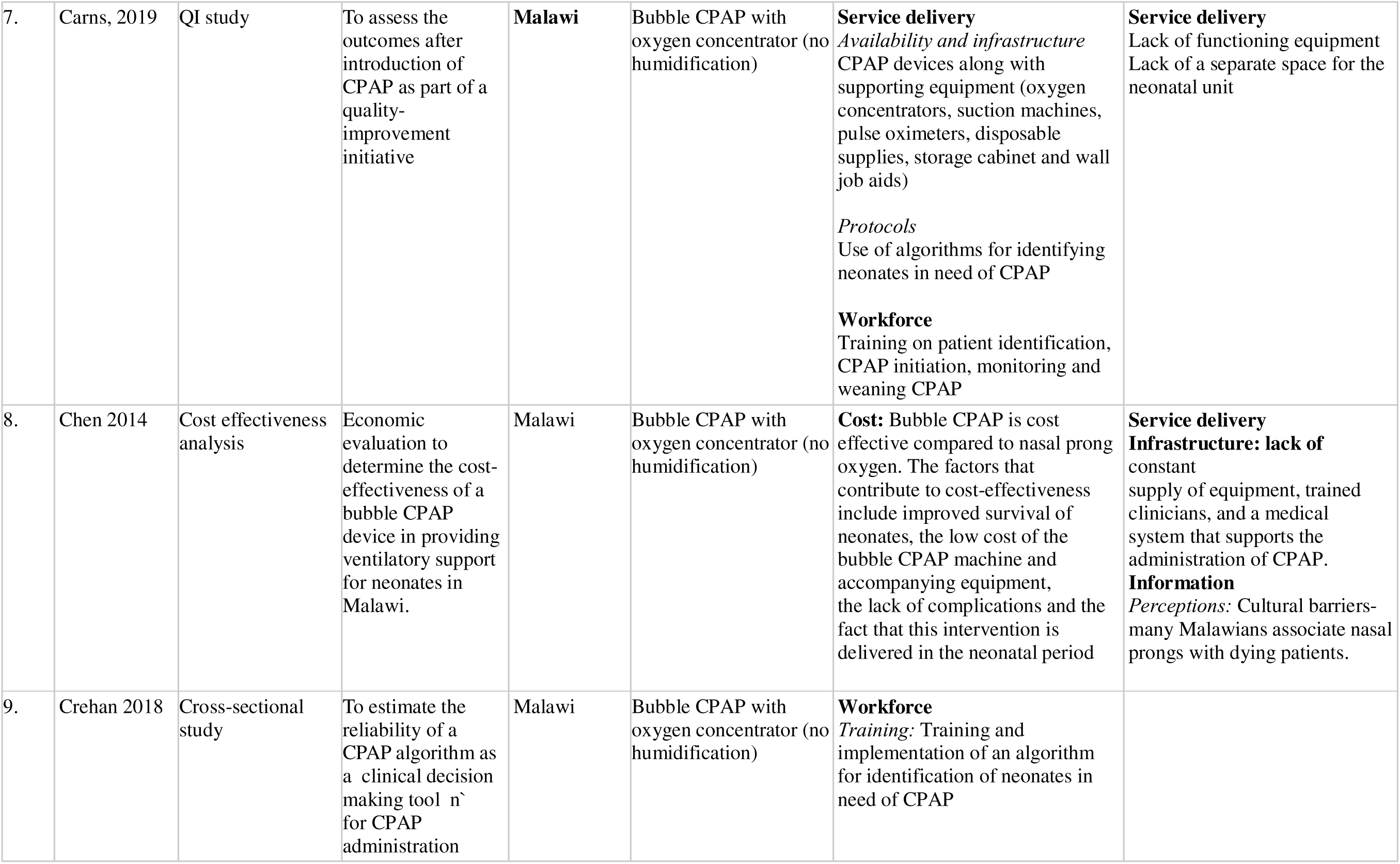
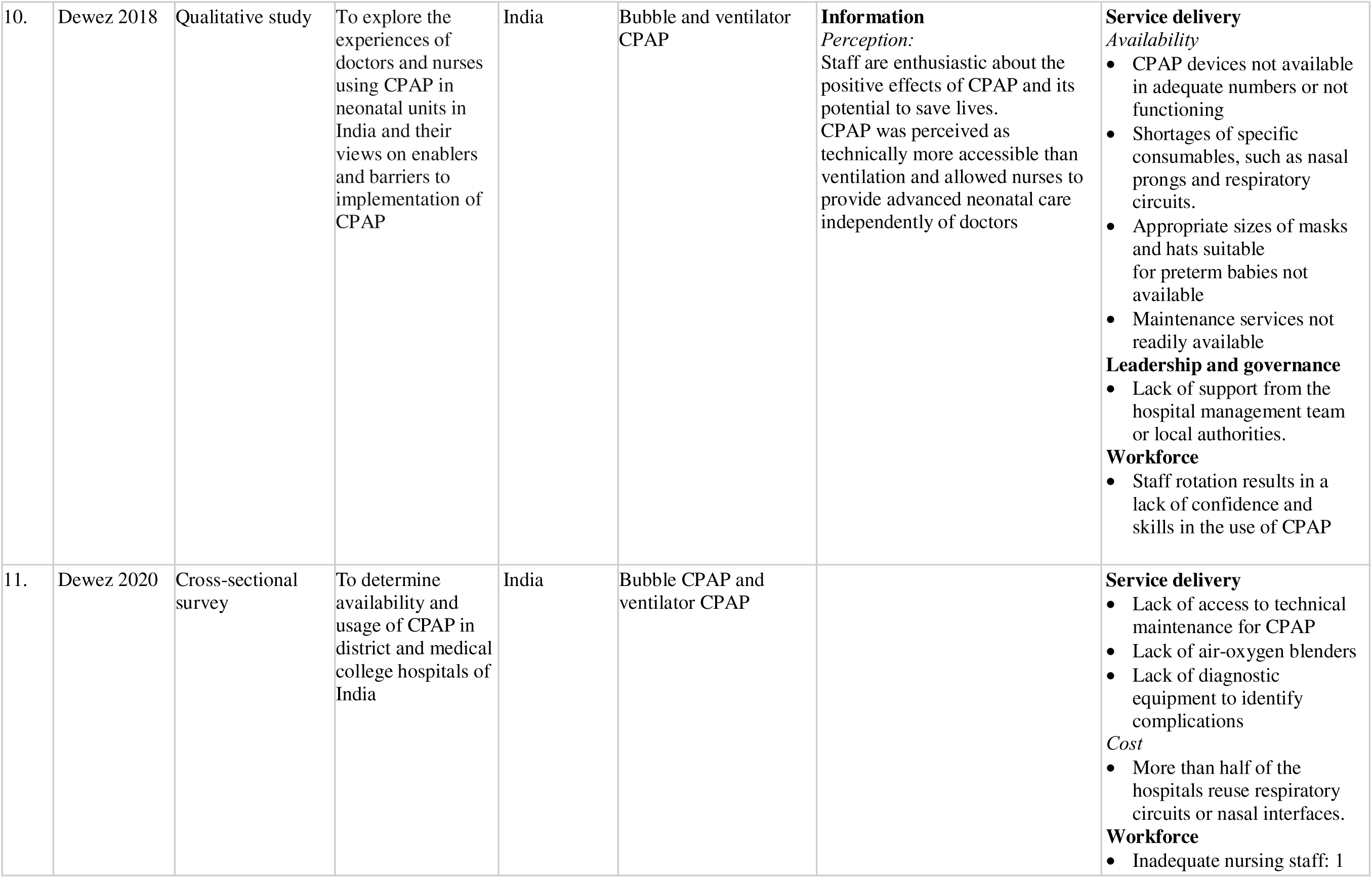
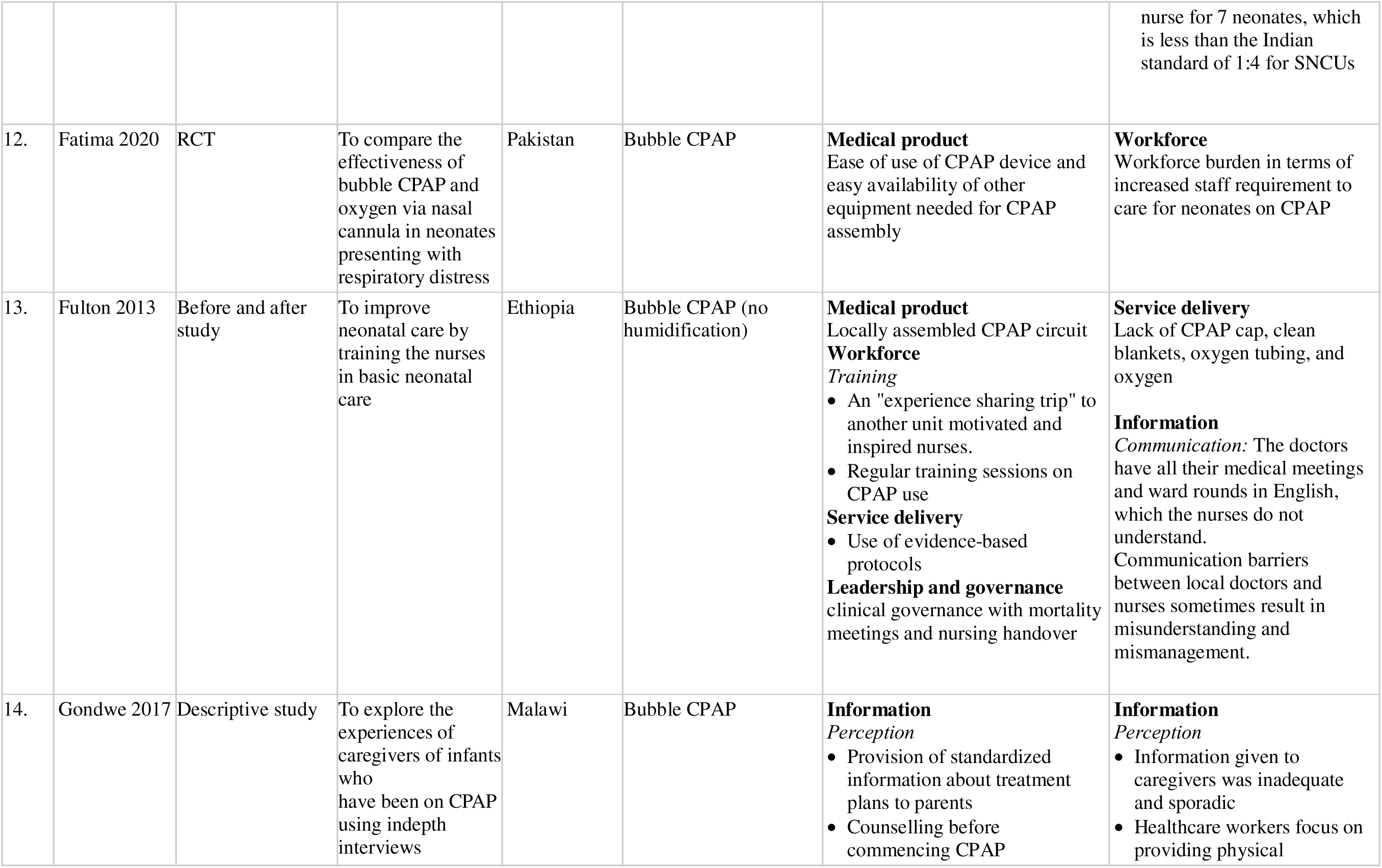
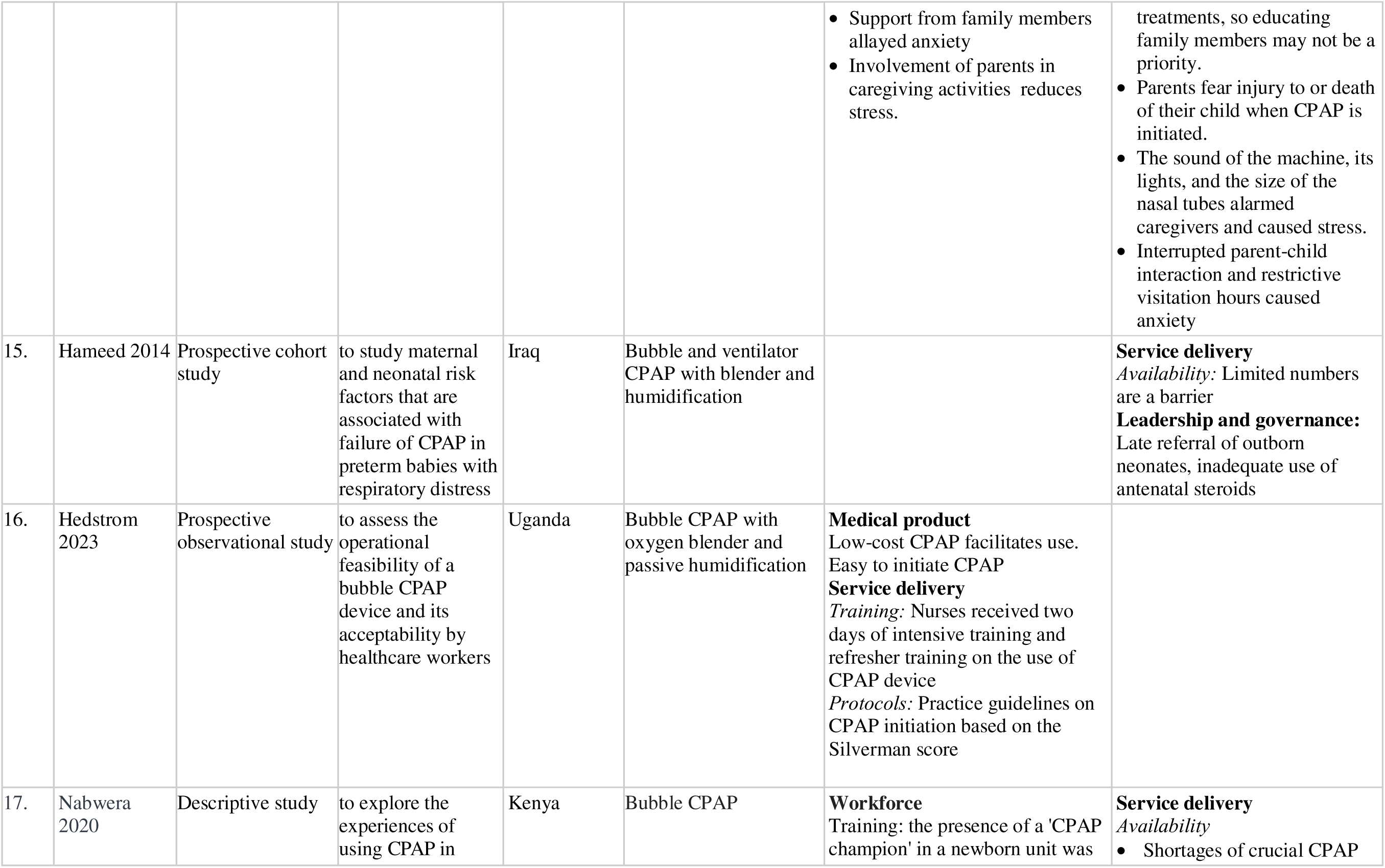
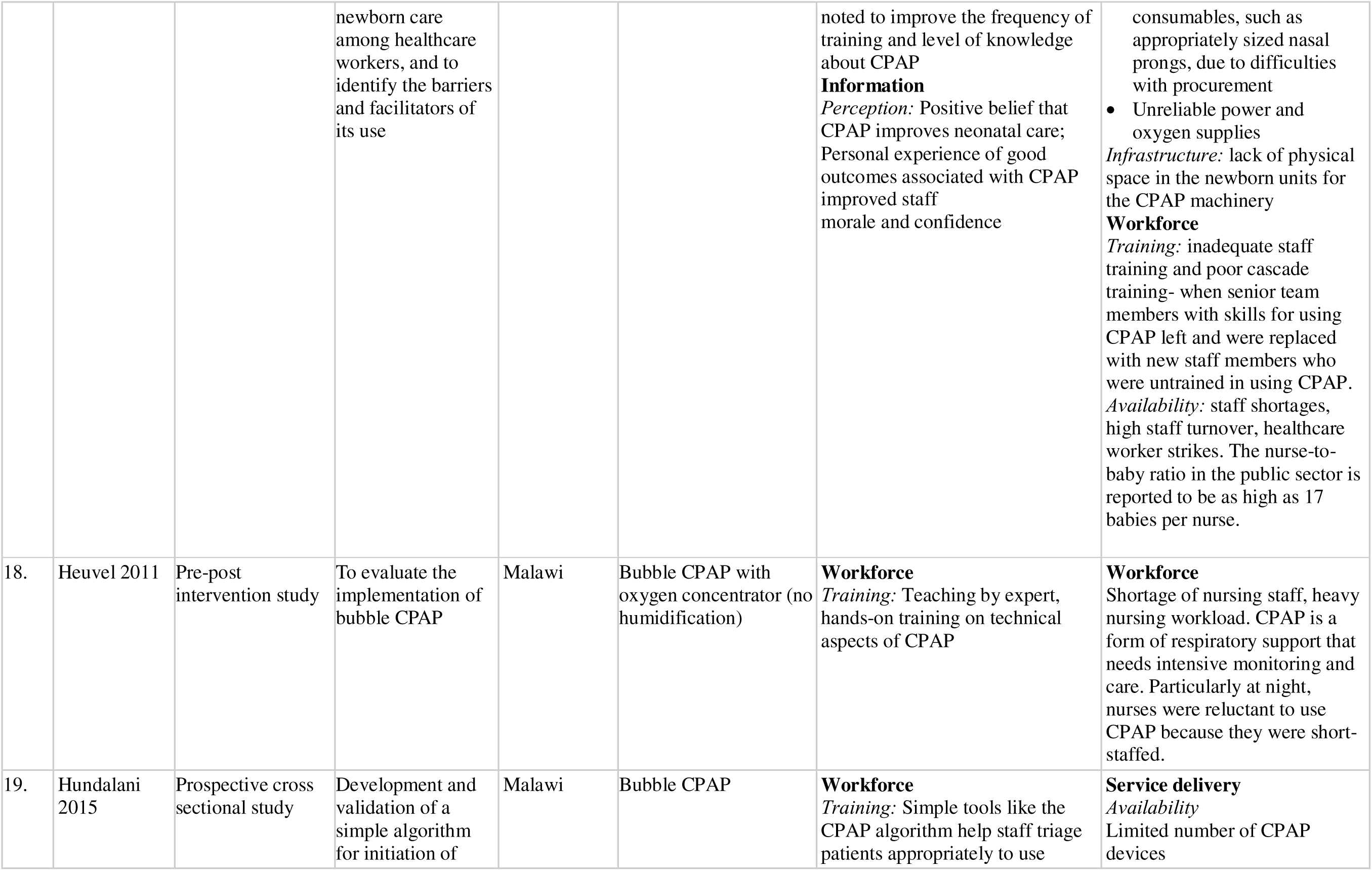
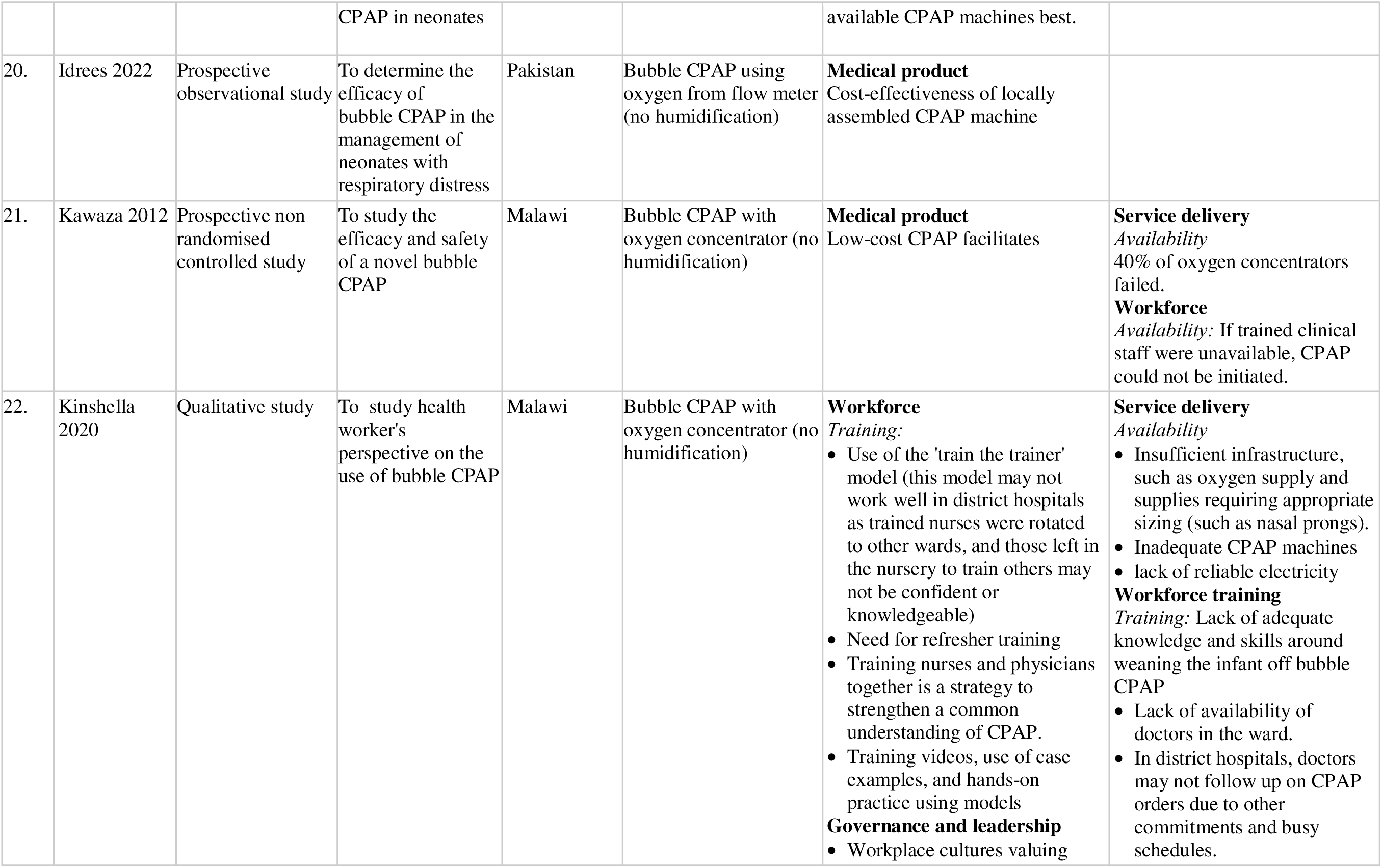
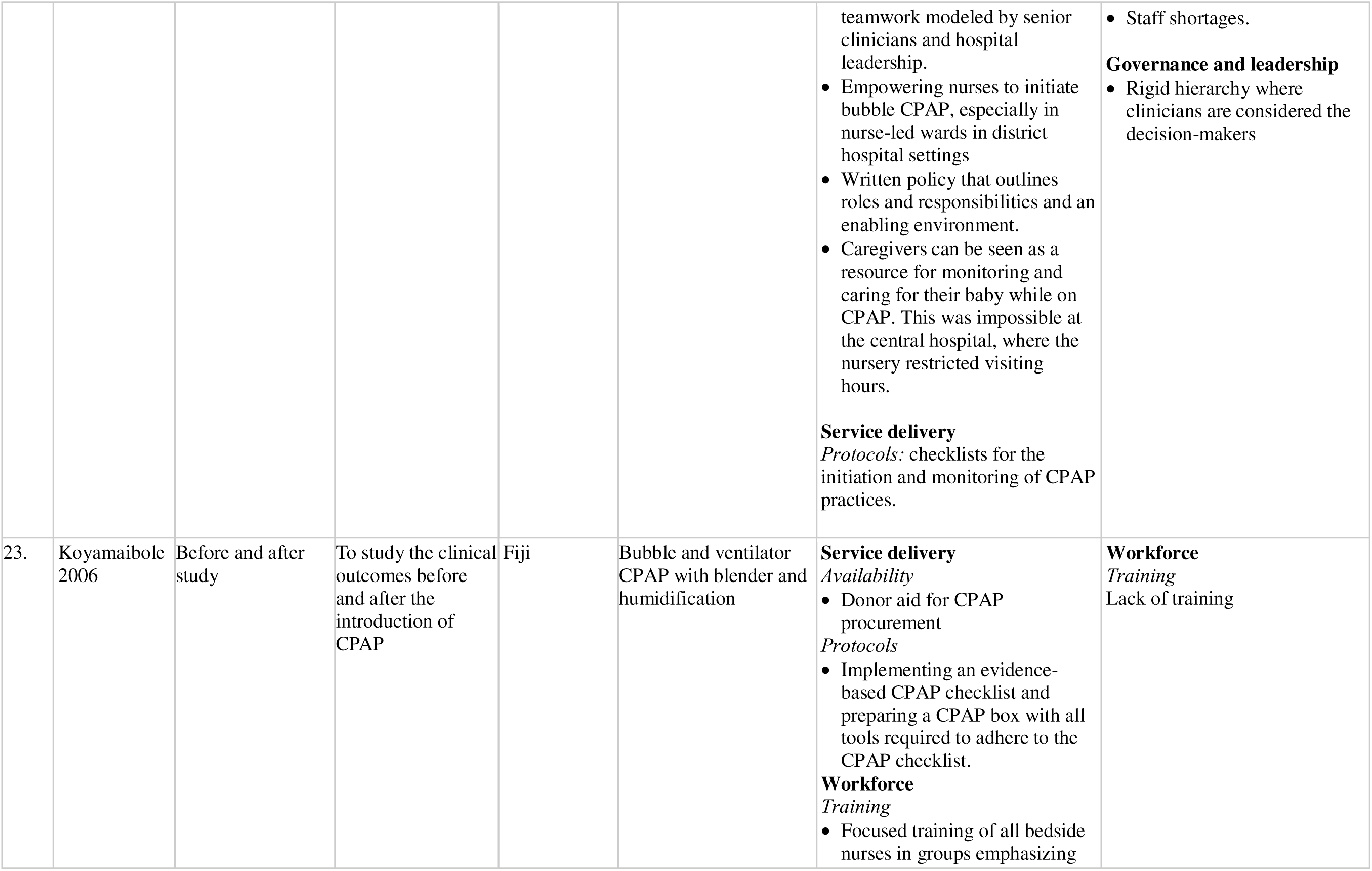
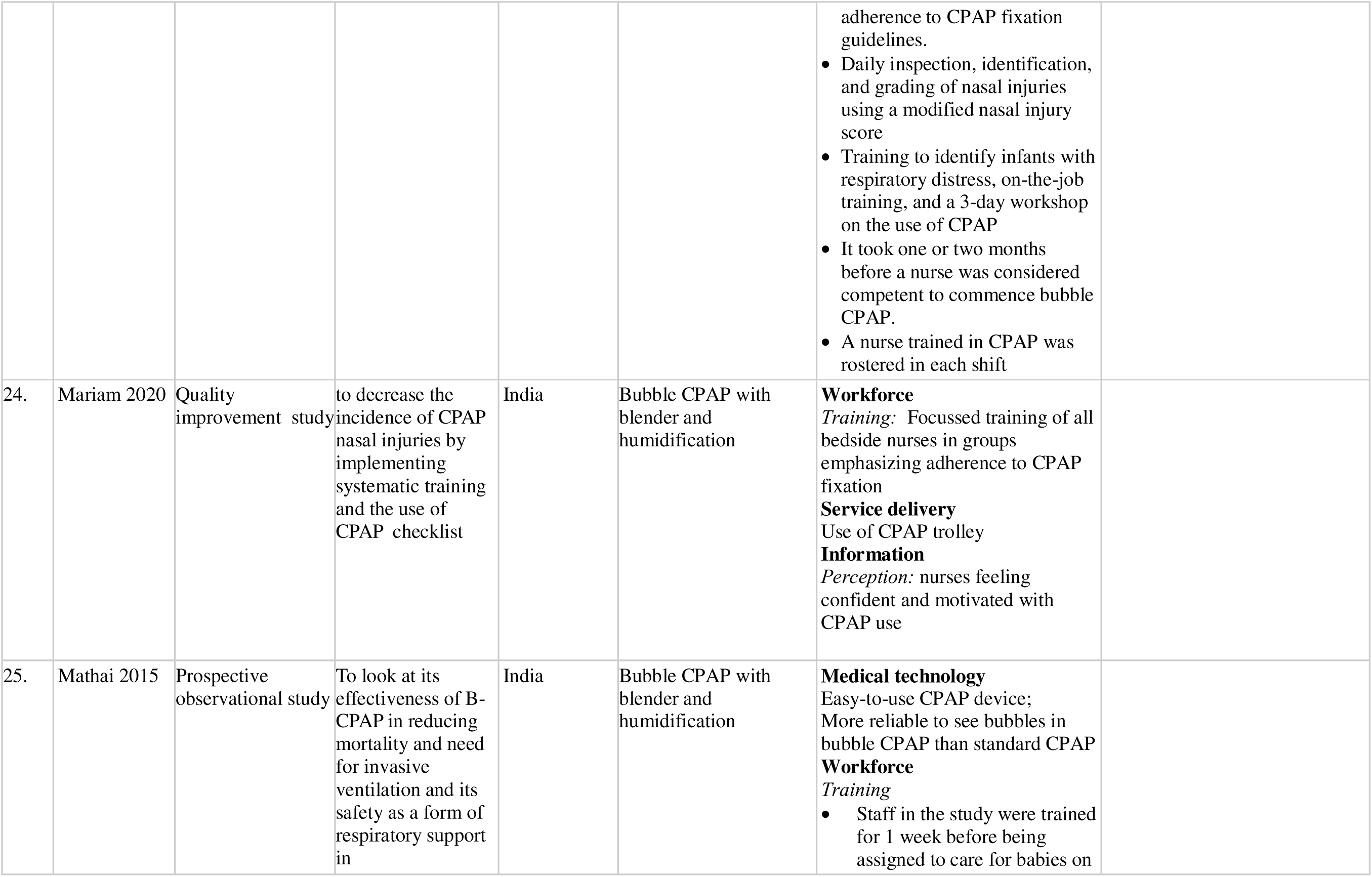
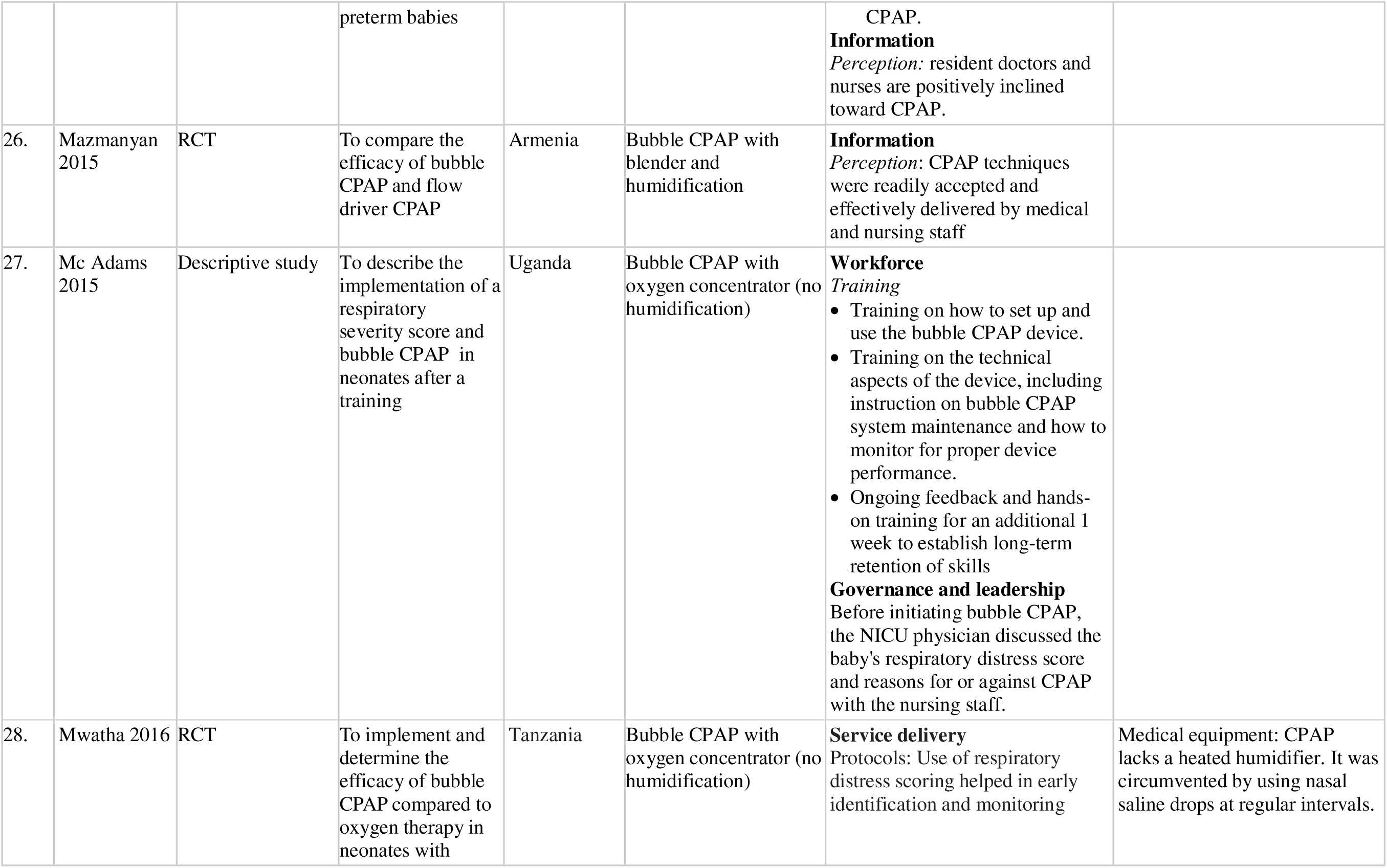
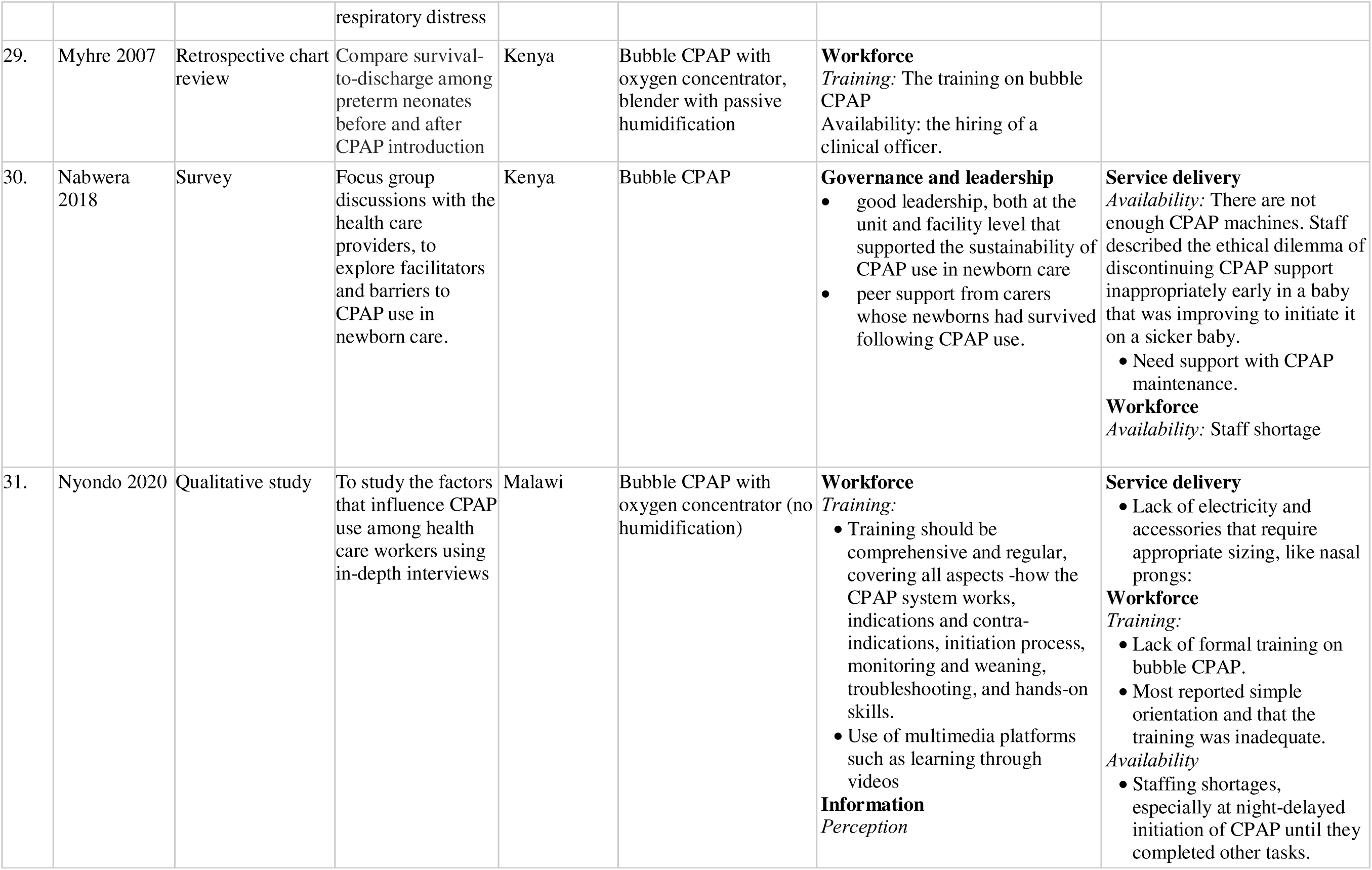
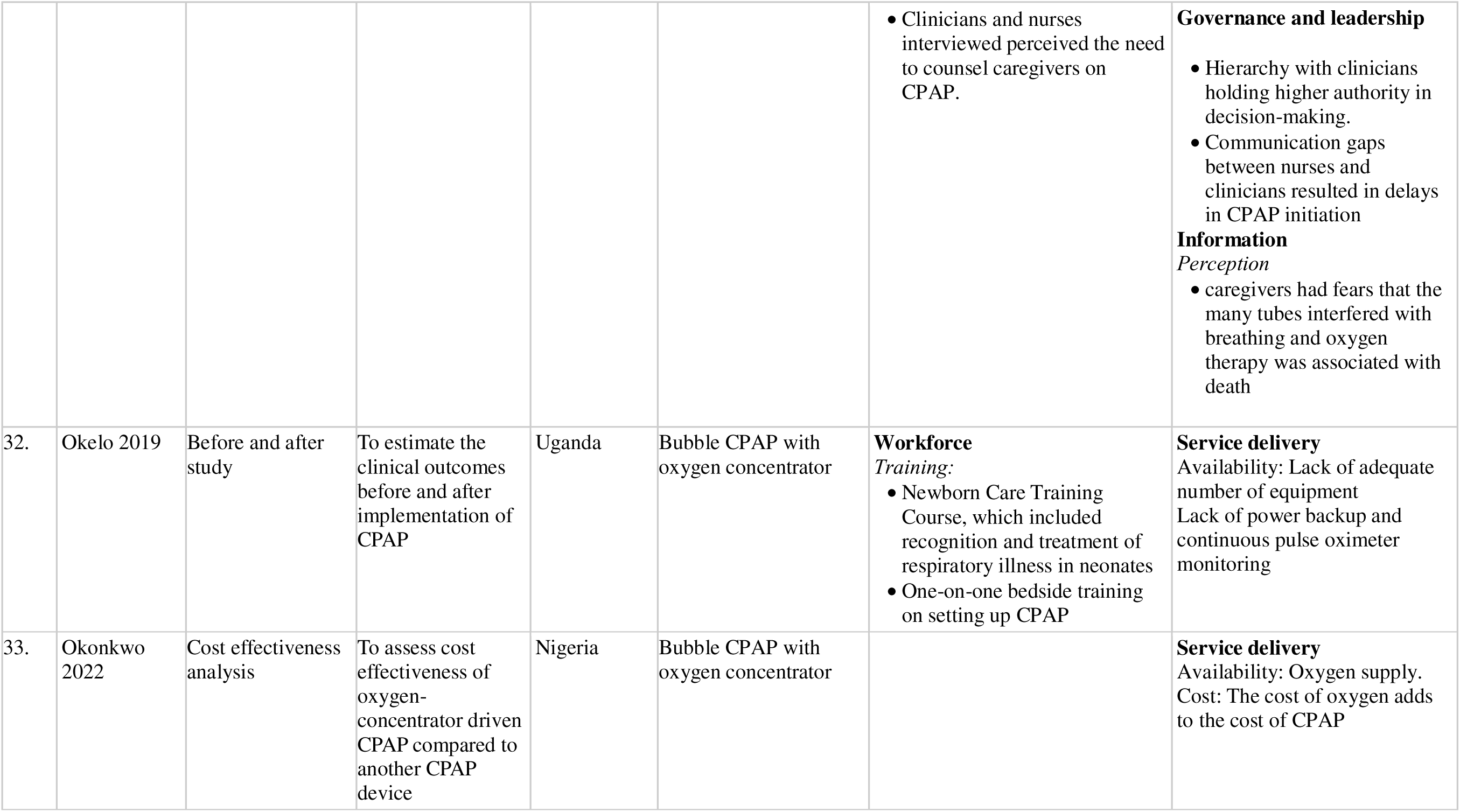
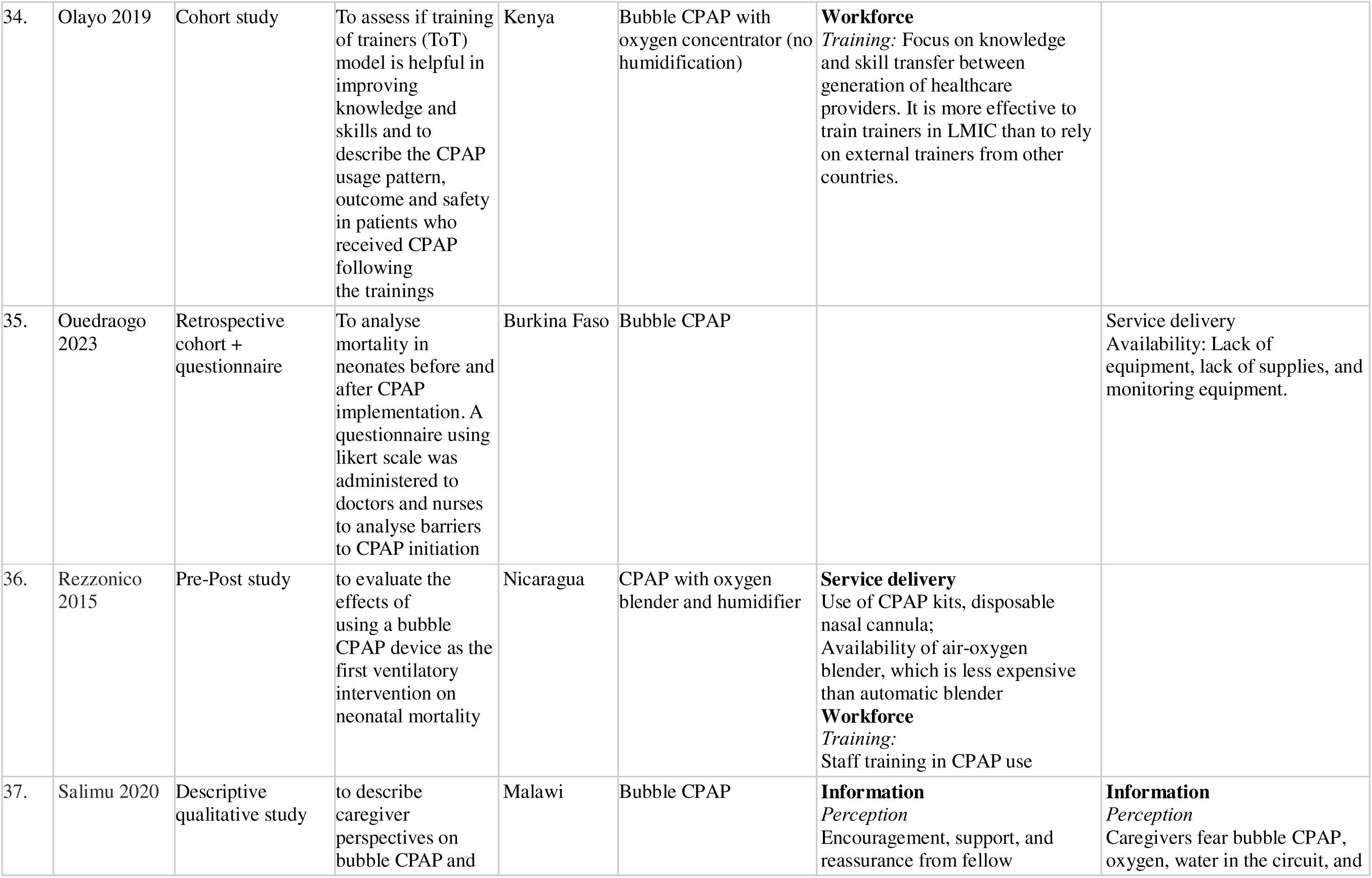
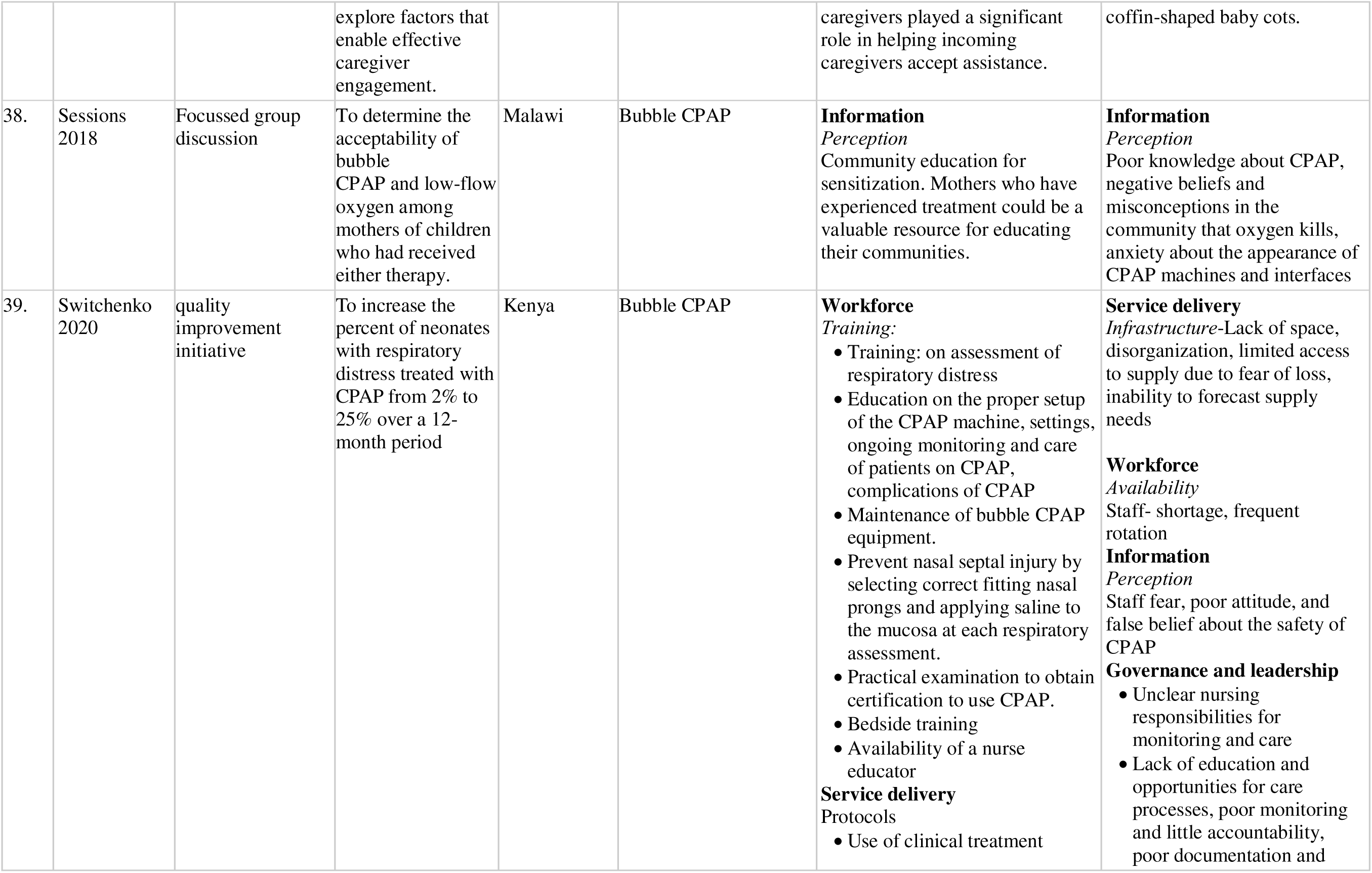
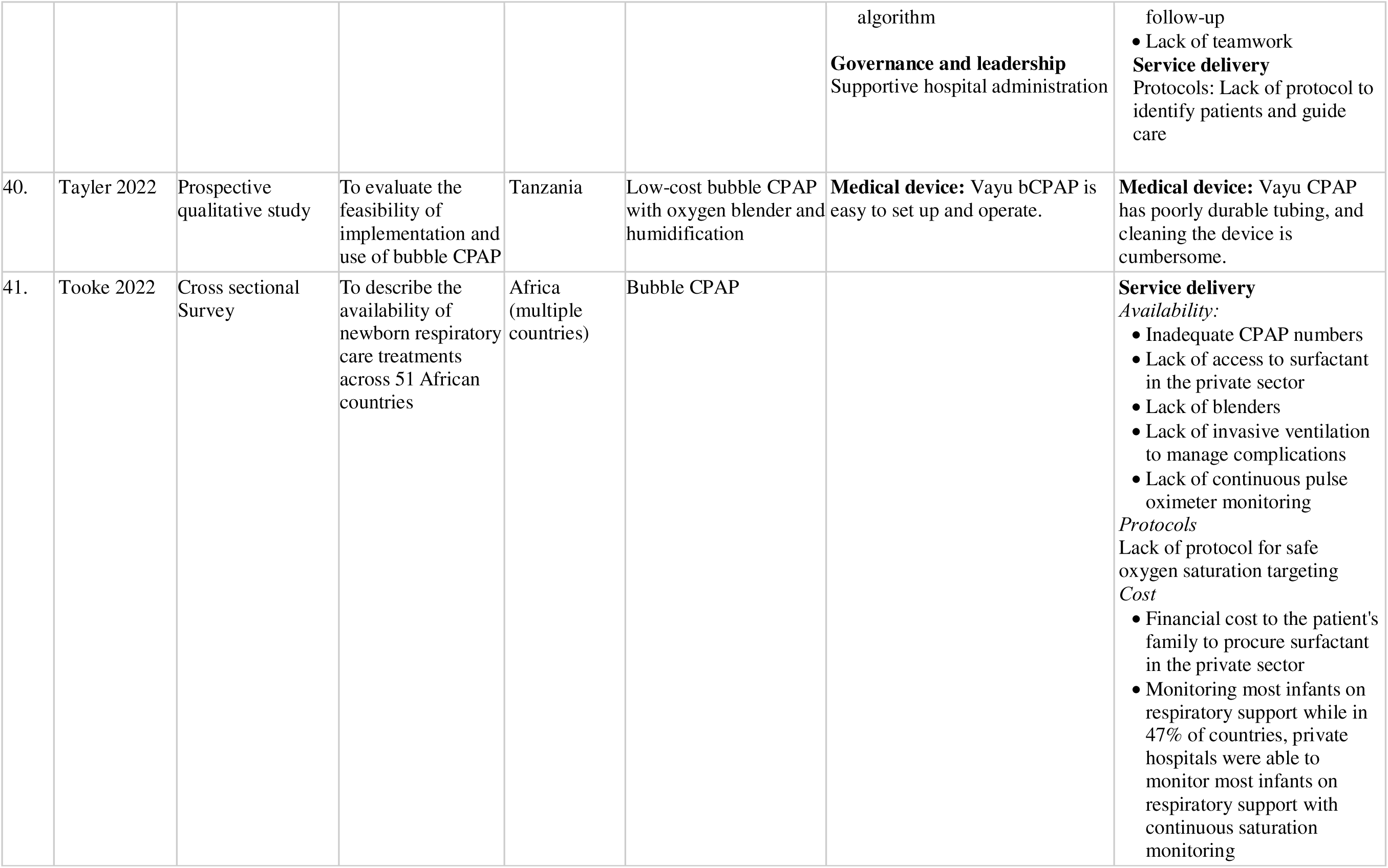
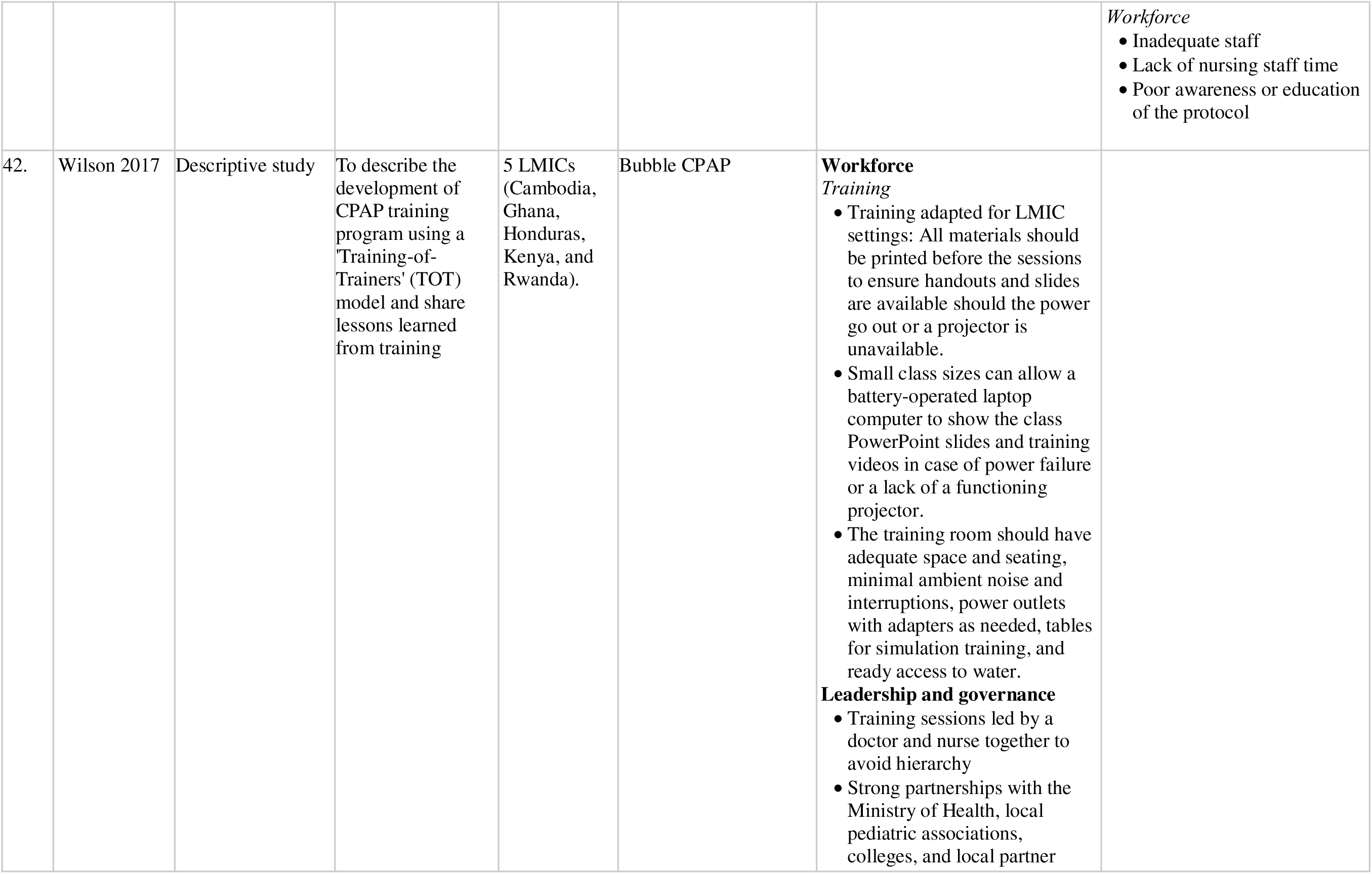
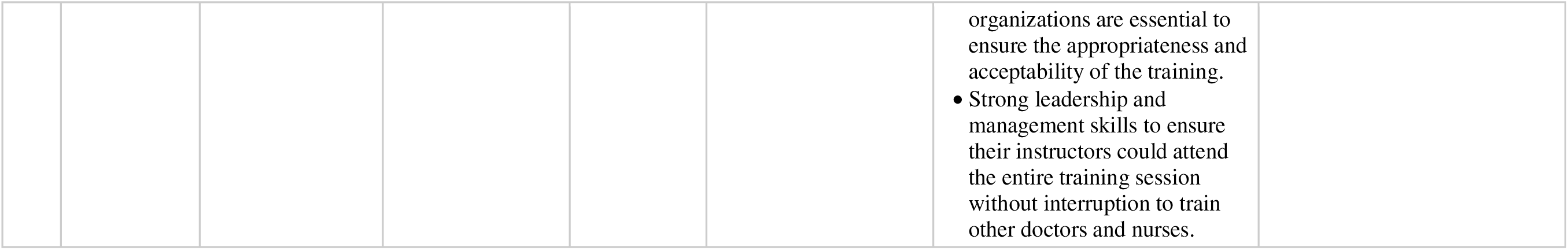
Facilitators and barriers reported in included studies.

### Facilitators for CPAP Implementation

Healthcare provider training (Figure 3) emerged as the most important facilitator for CPAP implementation, followed by the availability of a structured unit protocol and the ease of CPAP application.

**Figure 3:**
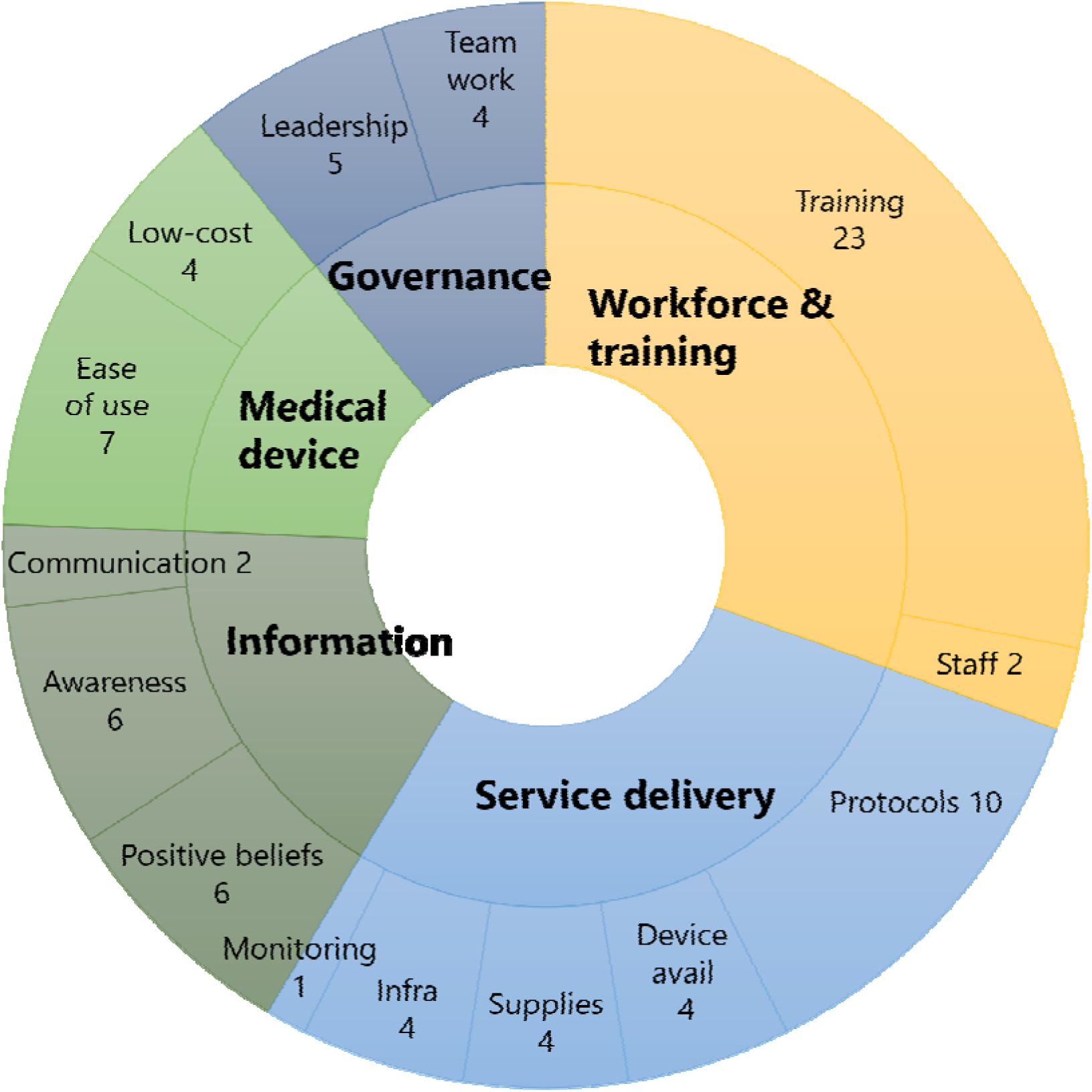
Ranking of facilitators of CPAP implementation. *The subthemes and the number of studies citing it are represented in this sunburst chart*.

#### 1. Medical equipment (CPAP device)

Ten studies reported that the intrinsic characteristics of bubble CPAP devices, such as ease of application and affordability, significantly facilitated their use. Bubble CPAP systems were noted to be easy to assemble without requiring additional expertise (8, 11, 19, 20, 23, 30, 45), allowing even junior nurses to set them up, thereby improving their confidence (17). Some models included built-in flow generators or venturi-type oxygen blenders (15, 33, 38), eliminating the need for compressed air sources or separate air–oxygen blenders.

Compared to ventilators and nasal prong oxygen, bubble CPAP devices are more affordable and cost-effective (8, 13, 14, 15, 23, 25, 26, 28, 45), making them especially suitable for public health systems.

#### 2. Service delivery

Fifteen studies reported facilitators within the service delivery theme. The application of evidence-based algorithms and checklists facilitated CPAP implementation (8, 20, 27, 28, 44). Standard tools, including the Silverman Anderson Score (23, 33), wall job aids (12), and innovative triaging algorithms for neonates experiencing respiratory distress (16, 24), aided healthcare providers in identifying infants who required CPAP.

Device availability (10, 14, 16, 30), whether through ward-strengthening programs (14) or scale-up initiatives, contributed to better implementation and survival outcomes (14).

Strengthening infrastructure, including the provision of CPAP devices, oxygen concentrators, suction machines, pulse oximeters, consumables, extra beds, and upgrades to plumbing and electrical facilities, has been shown to improve outcomes (12–14). An adequate supply of CPAP kits and interfaces facilitated implementation (12, 19, 21, 41) and was associated with reductions in mechanical ventilation rates and mortality (41).

#### 3. Workforce

Two studies reported that an increase in the healthcare workforce facilitated CPAP delivery (34, 44). Training of healthcare workers (HCWs) emerged as the most prominent enabler, as noted by 23 studies. Training methods included online video modules (9), peer mentorship for nurses (7, 10, 12, 27), instruction on early CPAP initiation algorithms (13, 14, 16, 24), train-the-trainer models (39, 48), hands-on workshops (20, 23, 28–30, 34, 36, 37, 41, 44), and periodic refresher training (23, 28, 44). Nine studies highlighted improved clinical outcomes linked to training, such as increased CPAP usage, fewer nasal injuries, a reduced need for mechanical ventilation, and improved survival rates (12, 14, 20, 23, 28–30, 47).

#### 4. Information

Twelve studies examined perceptions of CPAP among staff and parents (8, 11, 15, 20, 21, 31, 36, 42–44). Positive experiences and beliefs held by HCWs regarding CPAP led to improved clinical outcomes and care quality, as well as enhanced staff confidence and motivation (8, 17, 29–31, 35). Clinicians viewed CPAP as essential for preventing mechanical ventilation and its associated complications (17, 35).

Supportive elements, such as peer encouragement, parental involvement, interactions among caregivers, and family support, played significant roles in shaping positive perceptions (7, 21, 27, 36, 42, 43). Parental counseling and clear communication about the infant’s treatment reduced anxiety and facilitated acceptance of CPAP.

#### 5. Governance and leadership

Nine studies discussed facilitators related to governance and leadership, highlighting aspects such as leadership support and teamwork (7, 8, 12, 20, 27, 32, 35, 44, 48). Effective leadership within facilities enhanced the sustainability of CPAP use in neonates (7, 12, 27, 48). Collaborations with ministries of health, pediatric associations, and partner organizations increased the relevance and acceptance of training programs (44). Supportive hospital administration played a crucial role in sustainability by ensuring the maintenance of supplies and equipment (44).

A culture of teamwork, fostered by senior clinicians and leadership, created an enabling environment (27, 48). Nurses were empowered to initiate CPAP, especially in nurse-led district hospital wards. Joint training sessions for doctors and nurses reduced hierarchy and encouraged collaboration (8). The presence of a designated ‘CPAP champion’ in newborn units enhanced staff skills and knowledge (35). Written policies outlining staff roles and responsibilities, along with supportive environments, facilitated CPAP implementation.

### Barriers to CPAP implementation

The most critical barriers to CPAP implementation were a shortage of trained healthcare providers and limited availability of CPAP devices, followed by inadequate supplies and accessories, insufficient infrastructure, and a lack of training (Figure 4).

**Figure 4:**
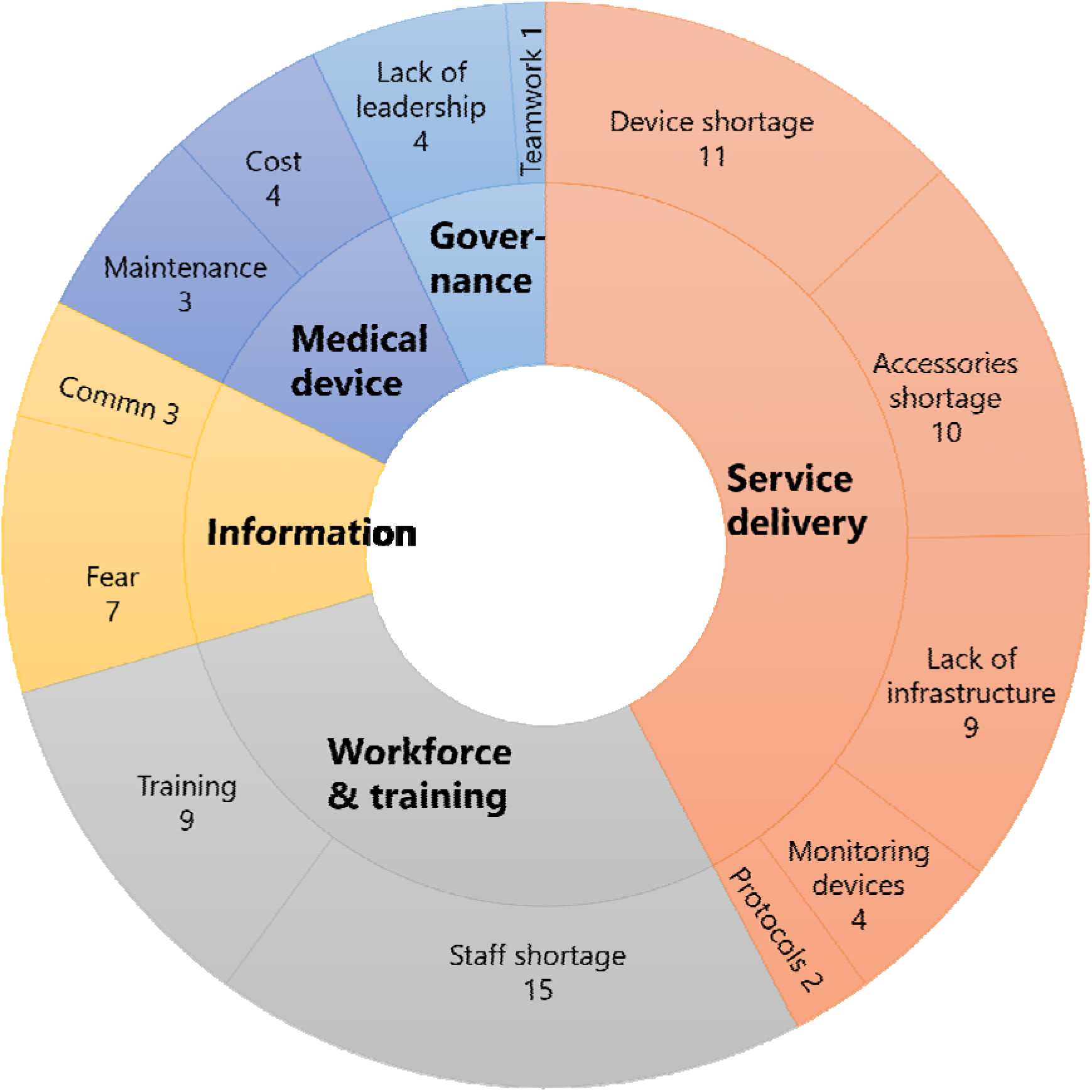
Ranking of barriers to CPAP implementation. *The subthemes and the number of studies citing it are represented in this sunburst chart*

#### 1. Medical devices

A multi-country survey of 49 African health facilities emphasized the need for equipment budgets that cover not only procurement but also maintenance, repair, and part replacement (46). The cost of oxygen, blenders, and CPAP circuits was a limiting factor in four studies (8, 11, 38, 46).

#### 2. Service delivery

Nineteen studies reported barriers related to service delivery, particularly the limited availability of CPAP devices (7, 12, 15, 17, 18, 22, 24, 35, 37, 40, 46). In a qualitative study, Nabwera et al. emphasized the insufficient number of devices as a significant barrier (35).

Frequent power outages, high fuel costs for generators, and unreliable electricity impeded device operation (8, 10, 27, 36, 37). In many settings, piped oxygen was unavailable, necessitating reliance on cylinders (8, 35). Additional barriers included space constraints, disorganized environments, limited access to supplies, and difficulties in forecasting supply needs (14, 44). Overcrowding in neonatal areas often resulted in multiple infants sharing limited space, complicating the initiation of CPAP (35).

The lack of accessories such as CPAP circuits and nasal interfaces was another significant obstacle (8, 11, 17, 18, 26, 27, 35–37, 46). Due to costs, tubing was often disinfected and reused (11, 17, 18). The scarcity of other essential equipment such as oxygen concentrators, humidifiers (26), blenders (46), invasive ventilation for managing CPAP failure (37, 46), and radiography for detecting air leaks (17) was also reported.

The lack of protocols for initiating, monitoring, and safely targeting oxygen levels restricted standardization (44, 46). Additionally, insufficient monitoring tools like pulse oximeters impeded the effective use of CPAP (17, 18, 40, 44). Surveys conducted in various hospitals revealed that the absence of oxygen saturation monitoring was a common barrier (17, 18, 44).

#### 3. Workforce and training

Sixteen studies identified gaps in human resources and a lack of training as major barriers. Inadequate staffing (7, 18, 26, 36, 47) and the frequent rotation of trained personnel resulted in shortages of skilled staff in neonatal units (9, 11, 17, 34). Training deficits were emphasized as significant obstacles to the initiation of CPAP (8, 10, 11, 26–28, 35, 36, 46). Challenges reported included short training durations, insufficient teaching materials, a lack of mentorship for new staff (8), limited hands-on practice (10), and ineffective cascade training following staff turnover (35).

#### 4. Information

Nine studies reported negative perceptions among caregivers and families that adversely affected CPAP uptake (11, 20, 21, 36, 42–44). Misconceptions about oxygen use, associating it with impending death, and fears about water in the CPAP circuit contributed to the reluctance (11, 15, 21, 36, 42–44). Language barriers between local doctors and nurses often led to miscommunication and mismanagement (20, 21, 36). Additionally, poor communication with families undermined their understanding and cooperation with CPAP therapy.

#### 5. Governance and leadership

Governance-related barriers were identified in five studies (17, 22, 27, 36, 44). These included insufficient support from hospital leadership or local authorities, rigid clinical hierarchies that allowed physicians to retain decision-making authority, and unclear role definitions for nurses. Poor interprofessional communication, minimal accountability, inadequate documentation, and limited teamwork contributed to delays in CPAP initiation (44).

## Discussion

This systematic review synthesizes evidence from 42 studies across 18 LMICs, providing a comprehensive overview of the facilitators and barriers to CPAP implementation. Three consistent enablers emerged: (1) structured unit-level protocols and clinical checklists that guided standardized care delivery; (2) the availability of user-friendly and low-maintenance CPAP devices suitable for use by mid-level providers; and (3) systematic training programs that built staff competence, motivation, and accountability. Barriers were multifactorial, encompassing infrastructure gaps, human resource constraints, and supply chain failures.

Most notably, shortages of trained staff, frequent rotation without mentoring, and a lack of consumables (e.g., nasal prongs, humidifiers, circuits) disrupted CPAP delivery. Device limitations, including lack of oxygen blenders and humidification, were also prominent, raising concerns about safety in resource-limited contexts. These findings underscore that CPAP implementation must be regarded as a systems intervention rather than a device-specific solution.

Our findings align with previous syntheses, including those by Kinshella et al. and Dada et al., which documented similar themes in sub-Saharan Africa. However, this review builds on those efforts by incorporating evidence from Asian LMICs and organizing the findings using the WHO health systems framework. While prior reviews emphasized affordability and device simplicity as key enablers, our findings underscore the equally critical role of context-specific training and operational challenges, such as hierarchical team structures, interdisciplinary gaps, and clinical governance, that can hinder implementation even in relatively well-resourced tertiary settings.

Notable regional differences emerged. In sub-Saharan Africa, foundational challenges such as intermittent electricity, unreliable oxygen supply, and a lack of physical space were frequently reported, especially in rural and district-level hospitals. Device availability was often addressed through local innovation or donations, but critical features like humidification and oxygen blending were frequently absent. In contrast, Asian LMICs reported fewer structural constraints but more often described financial barriers, particularly out-of-pocket costs for oxygen and consumables borne by families. These studies also emphasized challenges related to staff attrition and retention, underscoring the need for financial protection and workforce policies tailored to local realities. Such regional variation suggests a need for tailored implementation strategies: infrastructure and supply chain development in African settings and policy reforms regarding financing and workforce retention in Asia.

Facility-level differences further shaped implementation success. Tertiary hospitals in African countries faced organizational barriers, including fragmented team communication, unclear role distinctions between physicians and nurses, and resistance to interdisciplinary collaboration. In contrast, district hospitals struggled with more fundamental gaps such as limited or no formal training programs, rapid staff turnover, and environments unsuitable for skill retention. These challenges call for tier-specific strategies. Tertiary centers would benefit from investments in team-based care models, structured mentorship, and clinical leadership development. Conversely, district hospitals require foundational support: workforce expansion, basic infrastructure, and consumable logistics.

This review has several strengths. It is the first to incorporate studies from a wide geographic range across both African and Asian LMICs while synthesizing data from various study designs—spanning program evaluations to qualitative assessments. The thematic synthesis, aligned with WHO’s health system domains, enhances its relevance for policymakers and implementers. The application of rigorous methodology, including independent screening and dual data extraction, adds credibility to the findings. However, a few limitations should be noted. Most included studies were observational or qualitative, which limits causal inference. There was a geographic skew toward African countries, with limited representation from Latin America and fewer long-term outcome data. Additionally, cost-effectiveness, caregiver perspectives, and post-discharge follow-up were underreported, highlighting important gaps for future research.

## Conclusions

Improving CPAP implementation in LMICs requires a systems-based approach that goes beyond just device availability. Sustained success depends on a reliable infrastructure (electricity, oxygen), a consistent supply of consumables, motivated and trained staff, and clinical governance tools like protocols and checklists. Regional and facility-level variations necessitate differentiated strategies—investments in infrastructure and oxygen systems in African settings and financial protection and staff retention in Asia. CPAP scale-up should be integrated within resilient and responsive health systems. The findings from this review offer actionable guidance for policymakers and program designers seeking to address implementation gaps and accelerate progress toward global neonatal survival goals.

## Data Availability

All data produced in the present study are available upon reasonable request to the authors

## Notes

### Competing Interest Statement

The authors have declared no competing interest.

### Funding Statement

World Health Organization (WHO)

