## Appendix for "Factors Affecting the Implementation of Continuous Positive Airway Pressure (CPAP) in Low- and Middle-Income Countries: A Qualitative Evidence Synthesis": Appendix.docx

Table 1: Search terms

Table 2: Embase search

Table 3: Pubmed search

Table 4: CINAHL search

Table 5: EBSCO search

Table 6: Description of various themes and sub-themes mentioned in the review

Table 7 : Indexed ranking methodology

Table 8: Participant details of studies enrolling healthcare workers and caregivers

**Table 1: Search terms**

| Population | infant, newborn/ or infant, low birth weight/ or infant, small for gestational age/ or infant, very low birth weight/ or infant, postmature/ or infant, premature/  (infant or newborn or neonate or neonatal or premature or low birth weight or VLBW or LBW or infant* or neonat*).mp. not animals/ |
| --- | --- |
| Setting | “Developing Countries” OR developing countries* OR under developed countries* OR lmic* OR ((less developed OR low income OR lower income OR low and middle income OR low middle income OR resource poor OR low middle income OR resource constrained OR low resource OR limited resource* OR resource limited OR “Africa South of the Sahara” OR “Central America” OR “South America” OR “Latin America” OR “Caribbean Region” OR “Mexico” OR “Asia” OR “Asia, Central” OR “Asia, Northern” OR “Asia, Southeastern” OR “Asia, Western” OR OR “Korea” OR Afghanistan* OR Africa OR African[(tiab]) OR Algeria*[(tiab]) OR Angola*[(tiab]) OR Bangladesh*[(tiab]) OR Benin*[(tiab]) OR Bhutan*[(tiab]) OR Bolivia*[(tiab]) OR “Burkina Faso”[(tiab]) OR Burkinabe[(tiab]) OR Burundi*[(tiab]) OR Cambodia*[(tiab]) OR Cameroon*[(tiab]) OR “Cape Verde”[(tiab]) OR “Cape Verdean”[(tiab]) OR “Central African Republic”[(tiab]) OR Chad*[(tiab]) OR Comoros[(tiab]) OR Comorian[(tiab]) OR Congo[(tiab]) OR Congolese[(tiab]) OR “Côte d’Ivoire”[(tiab]) OR “Ivory Coast”[(tiab]) OR Ivorian[(tiab]) OR Djibouti*[(tiab]) OR Egypt*[(tiab]) OR “El Salvador”[(tiab]) OR Salvadorian[(tiab]) OR Guinean[(tiab]) OR Eritrea*[(tiab]) OR Ethiopia*[(tiab]) OR Eswatini *[(tiab]) OR Gambia*[(tiab]) OR Ghana[(tiab]) OR Ghanaian[(tiab]) OR Guinea-Bissau*[(tiab]) OR Haiti*[(tiab]) OR Hondura*[(tiab]) OR India[(tiab]) OR Indian[(tiab]) OR Indonesia*[(tiab]) OR Iran*[(tiab]) OR Kenya[(tiab]) OR Kenyan[(tiab]) OR Kiribati[(tiab]) OR Korea*[(tiab]) OR Kyrgy*[(tiab]) OR Laos[(tiab]) OR Laotian*[(tiab]) OR Lebanon[(tiab]) OR Lebanese[(tiab]) OR Lesotho[(tiab]) OR Liberia*[(tiab]) OR Madagasca*[(tiab]) OR Malawi*[(tiab]) OR Mali[(tiab]) OR Malian*[(tiab]) OR Mauritania*[(tiab]) OR Micronesia*[(tiab]) OR Mongolia*[(tiab]) OR Morocc*[(tiab]) OR Mozambique[(tiab]) OR Mozambican[(tiab]) OR Myanmar[(tiab]) OR Nepal*[(tiab]) OR Nicaragua*[(tiab]) OR Niger[(tiab]) OR Nigeria[(tiab]) OR Pakistan*[(tiab]) OR “Papua New Guinea”[(tiab]) OR Philippine*[(tiab]) OR Filipino*[(tiab]) OR Rwanda*[(tiab]) OR Samoa*[(tiab]) OR Sao Tome*[(tiab]) OR Principe[(tiab]) OR Senegal*[(tiab]) OR Sierra Leon*[(tiab]) OR Solomon Island*[(tiab]) OR Somali*[(tiab]) OR Sri Lanka*[(tiab]) OR Sudan*[(tiab]) OR South Sudan*[(tiab]) OR Syria*[(tiab]) OR Tajik*[(tiab]) OR Tanzania*[(tiab]) OR “Timor Leste”[(tiab]) OR Togo*[(tiab]) OR Tonga*[(tiab]) OR Tunisia*[(tiab]) OR Uganda*[(tiab]) OR Ukraine*[(tiab]) OR Uzbekistan*[(tiab)] OR Vanuat*[(tiab]) OR Venezuela*[(tiab]) OR Vietnam*[(tiab]) OR “West Bank”[(tiab]) OR Yemen*[(tiab]) OR Zambia*[(tiab]) OR Zimbabwe*) |
| Exposure | Continuous Positive Airway Pressure/ or respiration, artificial/ or positive-pressure respiration, bubble continuous positive airway pressure or bubble CPAP or BCPAP. |

**Table 2 : Embase search (as on July 2, 2023)**

| No. | Query | Results |
| --- | --- | --- |
| #20 | #13 AND #19 | 1277 |
| #19 | #14 OR #15 OR #16 OR #17 OR #18 | 7143333 |
| #18 | (albania OR algeria OR 'american samoa' OR argentina OR azerbaijan OR belarus OR belize OR bosnia OR herzegovina OR botswana OR brazil OR bulgaria OR china OR colombia OR 'costa rica or croatia' OR cuba OR dominica OR 'dominica republic' OR ecuador OR 'equatorial guinea' OR fiji OR gabon OR grenada OR guyana OR iran OR 'islamic rep' OR iraq OR jamaica OR kazakhstan OR lebanon OR libya OR macedonia OR fyr) AND malaysia OR maldives OR 'marshall islands' OR mauritius OR mexico OR montenegro OR namibia OR nauru OR panama OR paraguay OR peru OR romania OR 'russian federation' OR samoa OR serbia OR 'south africa' OR 'st. lucia' OR 'st. vincent' OR grenadines OR suriname OR thailand OR tonga OR turkey OR turkmenistan OR tuvalu OR venezuela | 2556300 |
| #17 | angola OR armenia OR bangladesh OR bhutan OR bolivia OR 'cabo verde' OR cambodia OR cameroon OR djibouti OR egypt OR 'el salvador' OR georgia OR ghana OR guatemala OR honduras OR india OR indonesia OR jordan OR kenya OR kiribati OR kosovo OR 'kyrgyz republic' OR 'lao pdr' OR lao OR lesotho OR mauritania OR micronesia OR 'fed sts' OR moldova OR mongolia OR morocco OR myanmar OR nicaragua OR nigeria OR pakistan OR 'papua new guinea' OR philippines OR 'sao tome' OR principe OR 'solomon islands' OR 'sri lanka' OR sudan OR swaziland OR 'syrian arab republic' OR syria OR tajikistan OR 'timor leste' OR tunisia OR ukraine OR uzbekistan OR vanuatu OR vietnam OR 'west bank gaza' OR yemen OR zambia | 2696889 |
| #16 | 'low income countries' OR lmic OR 'developing countries' OR 'undeveloped countries' OR 'under developed countries' OR 'south asia' OR 'south asian' OR asia OR 'middle income countries' OR 'resource limited' OR africa OR southeastern OR 'pacific islands' OR 'micronesia' OR 'middle east' OR 'south america' | 880128 |
| #15 | afghanistan OR benin OR 'burkina faso' OR burundi OR 'central african republic' OR guinea OR chad OR comoros OR 'congo dem rep' OR congo OR eritrea OR ethiopia OR gambia OR 'guinea bissau' OR haiti OR korea OR 'dem peoples rep' OR somalia OR liberia OR madagascar OR malawi OR mali OR mozambique OR nepal OR niger OR rwanda OR senegal OR 'sierra leone' OR 'south sudan' OR tanzania OR togo OR uganda OR zimbabwe | 1285846 |
| #14 | 'low and middle income countries' OR 'low and middle income' OR lmic OR developing | 963045 |
| #13 | #11 AND #12 | 5558 |
| #12 | (((((((((continuous AND positive AND airway AND pressure OR continuous) AND positive AND pressure OR continuous) AND positive AND airway AND pressure OR cpap OR continuous) AND distending AND airway AND pressure OR continuous) AND positive AND transpulmonary AND pressure OR continuous) AND transpulmonary AND pressure OR continuous) AND inflating AND pressure OR continuous) AND negative AND distending AND pressure OR continuous) AND negative AND pressure OR continuous) AND airway AND pressure | 25260 |
| #11 | neonate* OR newborn* OR preterm* OR premature OR 'low birth weight' OR lbw OR vlbw OR elbw OR 'low birth weights' OR 'low-birth-weight' OR 'low birthweights' OR infant* OR 'pre-terms' OR 'pre-term' OR 'small gestational age' OR sga | 1890833 |

**Table 3 : PubMed search July 2, 2023**

| Search number | Query | Results |
| --- | --- | --- |
| 10 | #8 AND #9 | 2,453 |
| 9 | #3 OR #4 OR #5 OR#6 OR #7 | 1,29,61,199 |
| 8 | #1 AND #2 | 4,637 |
| 7 | (Albania OR Algeria OR "American Samoa" OR Argentina OR Azerbaijan OR Belarus OR Belize OR Bosnia OR Herzegovina OR Botswana OR Brazil OR Bulgaria OR China OR Colombia OR "Costa Rica OR Croatia" OR Cuba OR Dominica OR "Dominica Republic" OR Ecuador OR "Equatorial Guinea" OR Fiji OR Gabon OR Grenada OR Guyana OR Iran OR "Islamic Rep" OR Iraq OR Jamaica OR Kazakhstan OR Lebanon OR Libya OR Macedonia OR FYR Malaysia OR Maldives OR "Marshall Islands" OR Mauritius OR Mexico OR Montenegro OR Namibia OR Nauru OR Panama OR Paraguay OR Peru OR Romania OR "Russian Federation" OR Samoa OR Serbia OR "South Africa" OR "St. Lucia" OR "St. Vincent" OR Grenadines OR Suriname OR Thailand OR Tonga OR Turkey OR Turkmenistan OR Tuvalu OR Venezuela).ab,ti. | 335 |
| 6 | (Angola OR Armenia OR Bangladesh OR Bhutan OR Bolivia OR "Cabo Verde" OR Cambodia OR Cameroon OR "Cote d’Ivoire" OR Djibouti OR Egypt OR "El Salvador" OR Georgia OR Ghana OR Guatemala OR Honduras OR India OR Indonesia OR Jordan OR Kenya OR Kiribati OR Kosovo OR "Kyrgyz Republic" OR "Lao PDR" OR Lao OR Lesotho OR Mauritania OR Micronesia OR "Fed Sts" OR Moldova OR Mongolia OR Morocco OR Myanmar OR Nicaragua OR Nigeria OR Pakistan OR "Papua New Guinea" OR Philippines OR "Sao Tome" OR Principe OR "Solomon Islands" OR "Sri Lanka" OR Sudan OR Swaziland OR "Syrian Arab Republic" OR Syria OR Tajikistan OR "Timor Leste" OR Tunisia OR Ukraine OR Uzbekistan OR Vanuatu OR Vietnam OR "West Bank Gaza" OR Yemen OR Zambia).ab,ti. | 120 |
| 5 | ("middle income countries" OR "low income countries" OR LMIC OR "developing countries" OR "undeveloped countries" OR "under developed countries" OR "south asia" OR "south Asian" OR Asia OR "middle income countries" OR "resource limited" OR Africa or Southeastern OR "Pacific Islands" OR "Micronesia" OR "Middle East" OR "South America") | 16,93,582 |
| 4 | (Afghanistan OR Benin OR "Burkina Faso" OR Burundi OR "Central African Republic" OR Guinea OR Chad OR Comoros OR "Congo Dem Rep" OR Congo OR Eritrea OR Ethiopia OR Gambia OR "Guinea Bissau" OR Haiti OR Korea OR "Dem Peoples Rep" OR Somalia OR Liberia OR Madagascar OR Malawi OR Mali OR Mozambique OR Nepal OR Niger OR Rwanda OR Senegal OR "Sierra Leone" OR "South Sudan" OR Tanzania OR Togo OR Uganda OR Zimbabwe).ab,ti. | 49 |
| 3 | "low and middle income countries" OR "low and middle income" OR LMIC OR developing | 62,04,742 |
| 2 | (continuous positive airway pressure[MeSH] OR continuous positive pressure OR continuous positive airway pressure OR CPAP OR continuous distending airway pressure OR continuous positive transpulmonary pressure OR continuous transpulmonary pressure OR continuous inflating pressure OR continuous negative distending pressure OR continuous negative pressure OR continuous airway pressure) | 30,218 |
| 1 | (((infant, newborn[MeSH] OR newborn*[TIAB] OR "new born"[TIAB] OR "new borns"[TIAB] OR "newly born"[TIAB] OR baby*[TIAB] OR babies*[TIAB] OR premature[TIAB] OR prematurity[TIAB] OR preterm[TIAB] OR "pre term"[TIAB] OR "low birth weight"[TIAB] OR "low birthweight"[TIAB] OR VLBW[TIAB] OR LBW[TIAB] OR infan*[TIAB] OR neonat*[TIAB]))) | 13,12,132 |

**Table 4: CINAHL search (July 2, 2023 at 00:02 )**

| **#** | **Query** | **Results** |
| --- | --- | --- |
| S15 | S5 AND S13 AND S14 | 160 |
| S14 | S1 OR S3 | 580,078 |
| S13 | S6 OR S7 OR S8 OR S9 OR S10 OR S11 OR S12 | 618,455 |
| S12 | (LMIC or LMICs or "third world" or "LAMI country" or "LAMI countries") | 9,901 |
| S11 | ((developing or "less* developed" or "under developed" or underdeveloped or "middle income" or "low* income") N4 (economy or economies)) | 278 |
| S10 | ((developing or "less* developed" or "under developed" or underdeveloped or "middle income" or "low* income" or underserved or "under served" or deprived or poor*) N4 (countr* or nation* or population* or world)) | 61,873 |
| S9 | (Africa or Asia or Caribbean or "West Indies" or "South America" or "Latin America" or "Central America") | 83,331 |
| S8 | Albania or Algeria or "American Samoa" or Argentina or Azerbaijan or Belarus or Belize or Bosnia or Herzegovina or Botswana or Brazil or Bulgaria or China or Colombia or "Costa Rica ORCroatia" or Cuba or Dominica or "Dominica Republic" or Ecuador or "Equatorial Guinea" or Fiji or Gabon or Grenada or Guyana or Iran or "Islamic Rep" or Iraq or Jamaica or Kazakhstan or Lebanon or Libya or Macedonia or FYR Malaysia or Maldives or "Marshall Islands" or Mauritius or Mexico or Montenegro or Namibia or Nauru or Panama or Paraguay or Peru or Romania or "Russian Federation" or Samoa or Serbia or "South Africa" or "St. Lucia" or "St. Vincent" or Grenadines or Suriname or Thailand or Tonga or Turkey or Turkmenistan or Tuvalu or Venezuela | 301,940 |
| S7 | Angola or Armenia or Bangladesh or Bhutan or Bolivia or "Cabo Verde" or Cambodia or Cameroon or "Cote d’Ivoire" or Djibouti or Egypt or "El Salvador" or Georgia or Ghana or Guatemala or Honduras or India or Indonesia or Jordan or Kenya or Kiribati or Kosovo or "Kyrgyz Republic" or "Lao PDR" or Lao or Lesotho or Mauritania or Micronesia or "Fed Sts" or Moldova or Mongolia or Morocco or Myanmar or Nicaragua or Nigeria or Pakistan or "Papua New Guinea" or Philippines or "Sao Tome" or Principe or "Solomon Islands" or "Sri Lanka" or Sudan or Swaziland or "Syrian Arab Republic" or syria or Tajikistan or "Timor Leste" or Tunisia or Ukraine or Uzbekistan or Vanuatu or Vietnam or "West Bank Gaza" or Yemen or Zambia | 177,988 |
| S6 | Afghanistan or Benin or "Burkina Faso" or Burundi or "Central African Republic" or Guinea or Chad or Comoros or "Congo Dem Rep" or Congo or Eritrea or Ethiopia or Gambia or "Guinea Bissau" or Haiti or Korea or "Dem Peoples Rep" or Somalia or Liberia or Madagascar or Malawi or Mali or Mozambique or Nepal or Niger or Rwanda or Senegal or "Sierra Leone" or "South Sudan" or Tanzania or Togo or Uganda or Zimbabwe | 93,677 |
| S5 | (continuous positive airway pressure OR continuous positive pressure OR CPAP OR continuous distending airway pressure OR continuous positive transpulmonary pressure OR continuous transpulmonary pressure OR continuous inflating pressure OR continuous negative distending pressure OR continuous negative pressure OR continuous airway pressure) | 8,002 |
| S4 | (continuous positive airway pressure OR continuous positive pressure OR CPAP OR continuous distending airway pressure OR continuous positive transpulmonary pressure OR continuous transpulmonary pressure OR continuous inflating pressure OR continuous negative distending pressure OR continuous negative pressure OR continuous airway pressure) | 8,002 |
| S3 | (infant or infants or infant’s or infantile or infancy or newborn* or "new born" or "new borns" or "newly born" or neonat* or baby* or babies or premature or prematures or prematurity or preterm or preterms or "pre term" or premies or "low birth weight" or "low birthweight" or VLBW or LBW) | 568,765 |
| S2 | TI ( (Neonate* or Newborn* or Preterm* or term or premature or "Low birth weight" or lbw or vlbw or elbw or "Low birth weights" or "Low birthweight" or "Low birthweights" or Infant* or "pre-terms" or "Pre-term" or "Small gestational age" or SGA or "Extremely premature") ) OR AB ( (Neonate* or Newborn* or Preterm* or term or premature or "Low birth weight" or lbw or vlbw or elbw or "Low birth weights" or "Low birthweight" or "Low birthweights" or Infant* or "pre-terms" or "Pre-term" or "Small gestational age" or SGA or "Extremely premature") ) | 614,340 |
| S1 | (MH "Infant+") OR (MH "Infant, Premature") OR (MH "Infant, Postmature") OR (MH "Infant, High Risk") OR (MH "Infant, Very Low Birth Weight") OR (MH "Infant, Small for Gestational Age") OR (MH "Infant, Large for Gestational Age") OR (MH "Infant, Newborn, Diseases+") | 300,431 |

**T**

**Table 5: EBSCOhost Research Databases**

| **#** | **Query** | **Results** |
| --- | --- | --- |
| S4 | S1 AND S2 AND S3 | 1,978 |
| S3 | ( Afghanistan or Benin or "Burkina Faso" or Burundi or "Central African Republic" or Guinea or Chad or Comoros or "Congo Dem Rep" or Congo or Eritrea or Ethiopia or Gambia or "Guinea Bissau" or Haiti or Korea or "Dem Peoples Rep" or Somalia or Liberia or Madagascar or Malawi or Mali or Mozambique or Nepal or Niger or Rwanda or Senegal or "Sierra Leone" or "South Sudan" or Tanzania or Togo or Uganda or Zimbabwe ) OR ( Angola or Armenia or Bangladesh or Bhutan or Bolivia or "Cabo Verde" or Cambodia or Cameroon or "Cote d’Ivoire" or Djibouti or Egypt or "El Salvador" or Georgia or Ghana or Guatemala or Honduras or India or Indonesia or Jordan or Kenya or Kiribati or Kosovo or "Kyrgyz Republic" or "Lao PDR" or Lao or Lesotho or Mauritania or Micronesia or "Fed Sts" or Moldova or Mongolia or Morocco or Myanmar or Nicaragua or Nigeria or Pakistan or "Papua New Guinea" or Philippines or "Sao Tome" or Principe or "Solomon Islands" or "Sri Lanka" or Sudan or Swaziland or "Syrian Arab Republic" or syria or Tajikistan or "Timor Leste" or Tunisia or Ukraine or Uzbekistan or Vanuatu or Vietnam or "West Bank Gaza" or Yemen or Zambia ) OR ( Albania or Algeria or "American Samoa" or Argentina or Azerbaijan or Belarus or Belize or Bosnia or Herzegovina or Botswana or Brazil or Bulgaria or China or Colombia or "Costa Rica ORCroatia" or Cuba or Dominica or "Dominica Republic" or Ecuador or "Equatorial Guinea" or Fiji or Gabon or Grenada or Guyana or Iran or "Islamic Rep" or Iraq or Jamaica or Kazakhstan or Lebanon or Libya or Macedonia or FYR Malaysia or Maldives or "Marshall Islands" or Mauritius or Mexico or Montenegro or Namibia or Nauru or Panama or Paraguay or Peru or Romania or "Russian Federation" or Samoa or Serbia or "South Africa" or "St. Lucia" or "St. Vincent" or Grenadines or Suriname or Thailand or Tonga or Turkey or Turkmenistan or Tuvalu or Venezuela ) OR ( (Africa or Asia or Caribbean or "West Indies" or "South America" or "Latin America" or "Central America") ) OR ( ((developing or "less* developed" or "under developed" or underdeveloped or "middle income" or "low* income" or underserved or "under served" or deprived or poor*) N4 (countr* or nation* or population* or world)) ) OR ( ((developing or "less* developed" or "under developed" or underdeveloped or "middle income" or "low* income") N4 (economy or economies)) ) OR ( (LMIC or LMICs or "third world" or "LAMI country" or "LAMI countries") ) | 15,936,708 |
| S2 | (continuous positive airway pressure OR continuous positive pressure OR CPAP OR continuous distending airway pressure OR continuous positive transpulmonary pressure OR continuous transpulmonary pressure OR continuous inflating pressure OR continuous negative distending pressure OR continuous negative pressure OR continuous airway pressure) | 39,180 |
| S1 | ( (MH "Infant+") OR (MH "Infant, Premature") OR (MH "Infant, Postmature") OR (MH "Infant, High Risk") OR (MH "Infant, Very Low Birth Weight") OR (MH "Infant, Small for Gestational Age") OR (MH "Infant, Large for Gestational Age") OR (MH "Infant, Newborn, Diseases+") ) OR ( (infant or infants or infant’s or infantile or infancy or newborn* or "new born" or "new borns" or "newly born" or neonat* or baby* or babies or premature or prematures or prematurity or preterm or preterms or "pre term" or premies or "low birth weight" or "low birthweight" or VLBW or LBW) ) | 3,981,979 |

**Table 6: Description of various themes and sub-themes mentioned in the review**

| Theme | Sub-theme | Explanation |
| --- | --- | --- |
| Medical device | Ease of application  Cost  Maintenance | The ease with which the device can be assembled and accessories fixed, determined the ease of use  Included the cost of CPAP device as well as cost of oxygen  Maintenance included equipment malfunction costs, replacement of device parts, and consumables |
| Service delivery | Device availability  Infrastructure  Protocols  Supportive facilities  Monitoring devices  Supplies and accessories | Availability of functioning equipment  Infrastructure included availability of adequate floor space for patient care and equipment, electricity, facility for oxygen, power supply, beds, water supply  Availability of protocols for initiating, monitoring and troubleshooting in CPAP  Oxygen blenders, humidifiers, surfactant therapy, facility for mechanical ventilation, and X-ray machines  Pulse oximetry, ROP screening  Availability of circuit tubings, caps, CPAP interfaces and tubings |
| Workforce and training | Staff availability  Training | Availability of staff, staff rotation, staff turnover  Online, hands-on, peer-mentorship and hybrid training on: CPAP initiation, maintenance, monitoring, troubleshooting and weaning. |
| Information | Beliefs  Family and community awareness  Communication | Health care workers’ perceptions about CPAP use  Parental perceptions about CPAP use  Communication between nurses and doctors; communication of healthcare workers with parents |
| Governance and leadership | Leadership  Teamwork | Hierarchy, local governance, responsibility, accountability and role allocation  Doctor-nurse relations, enabling environment |
